# Dissection of medical AI reasoning processes via physician and generative-AI collaboration

**DOI:** 10.1101/2023.05.12.23289878

**Authors:** Alex J. DeGrave, Zhuo Ran Cai, Joseph D. Janizek, Roxana Daneshjou, Su-In Lee

**Author notes:** indicates co-senior authorship (Roxana Daneshjou, Su-In Lee).

## Abstract

Despite the proliferation and clinical deployment of artificial intelligence (AI)-based medical software devices, most remain black boxes that are uninterpretable to key stakeholders including patients, physicians, and even the developers of the devices. Here, we present a general model auditing framework that combines insights from medical experts with a highly expressive form of explainable AI that leverages generative models, to understand the reasoning processes of AI devices. We then apply this framework to generate the first thorough, medically interpretable picture of the reasoning processes of machine-learning–based medical image AI. In our synergistic framework, a generative model first renders “counterfactual” medical images, which in essence visually represent the reasoning process of a medical AI device, and then physicians translate these counterfactual images to medically meaningful features. As our use case, we audit five high-profile AI devices in dermatology, an area of particular interest since dermatology AI devices are beginning to achieve deployment globally. We reveal how dermatology AI devices rely both on features used by human dermatologists, such as lesional pigmentation patterns, as well as multiple, previously unreported, potentially undesirable features, such as background skin texture and image color balance. Our study also sets a precedent for the rigorous application of explainable AI to understand AI in any specialized domain and provides a means for practitioners, clinicians, and regulators to uncloak AI’s powerful but previously enigmatic reasoning processes in a medically understandable way.

## Introduction

Medical artificial intelligence (AI) devices have proliferated in recent years^1^, but currently, the scientific and medical community poorly understands what factors influence AI outputs and whether these factors could lead to failures and harm to patients when AI is deployed in practice. The reasoning processes of these high-stakes devices—namely those that rely on neural networks and other complex “machine-learning” techniques, which automatically learn statistical patterns in large datasets—remain opaque to all stakeholders, including patients, medical providers, regulators, and even the developers of these AI systems. In principle, a detailed understanding of the reasoning processes of these AI devices could help us predict and prevent AI failures, help us improve AI models, and offer scientific value by contributing to the community’s knowledge of AI reasoning processes or their underlying training data. However, to our knowledge, no thorough medically interpretable picture of the reasoning process of a machine-learning–based medical image AI device yet exists. Prior efforts provide extremely limited *peeks* at medical AI reasoning processes^2, 3^, typically via techniques that “sanity check” whether a model is looking in the correct place^4–7^, and both these and more expressive techniques^8, 9^ typically suffer from lack of principled, medically informed analysis, precluding a thorough understanding. Indeed, despite technical developments in these explainable AI (XAI) tools, the gap between XAI tool output and pragmatic understanding of an AI device, particularly for image analysis and other “representation learning” AI systems, remains so large that efforts to apply XAI often miss severe faults in an AI device’s logic^10–13^, such as strong dependence on spurious “shortcut” features^4, 14^.

In exploring the reasoning processes of medical image AI, dermatology AI devices serve as a particularly impactful use case, for several reasons: numerous academic papers report high performance^15–17^; the first handful of companies have received CE approval to deploy their AI devices on patients in the European Economic Area^18, 19^; and multiple developers are working on approval from the United States Food and Drug Administration^20^. Dermatology AI devices, often targeted directly at consumers, may pose particular risks due to the lack of involvement from healthcare providers, potential for bias on skin tone^21^ and other sensitive attributes, and heterogeneity of user-acquired images, since there are no implemented DICOM standards in dermatology. Simultaneously, the *de facto* standard^5^ XAI modality to analyze image models—saliency maps, which highlight the regions of an image that most influence a model’s prediction—appear poorly suited to understand dermatology AI devices, which may be best explained in terms of dermatological concepts (e.g., “multiple colors of pigment”, “atypical pigment networks”) that spatially overlap or manifest diffusely throughout an image (Supplementary Fig. 1). Explanation of even a single prediction involves simultaneously high levels of technical AI knowledge and dermatology expertise, impeding a global understanding of the AI device’s behavior.

Here, we scrutinize numerous high-profile dermatology AI models to obtain the first thorough, medically interpretable picture of medical image AI reasoning processes. In the process, we showcase our workflow that combines explainable AI with human domain expertise (Fig. 1a). We demonstrate solutions to severe practical issues with explainable AI in the imaging domain, including (i) conceptualizing AI behavior in medically meaningful terms, (ii) addressing sampling challenges to form robust conclusions, and (iii) scaling from explanations of individual predictions to a global understanding of an AI device’s reasoning processes. At a high level, our workflow involves synthesis of counterfactual images, which answer the question “how might a given image plausibly differ to have elicited a different prediction from the AI?”, via generative models, which circumvent limitations of the *de facto* standard XAI modality (saliency maps) in medical image analysis. Our workflow continues with the analysis of thousands of such counterfactual images by dermatology experts, to characterize an AI device in human-understandable medical terms. Throughout the process, we emphasize rigor by mitigating problems of sampling and bias, via examination of numerous images, consideration of multiple datasets, and solicitation of insights independently from two dermatologists via a randomized and blinded analysis.

**Fig. 1.**
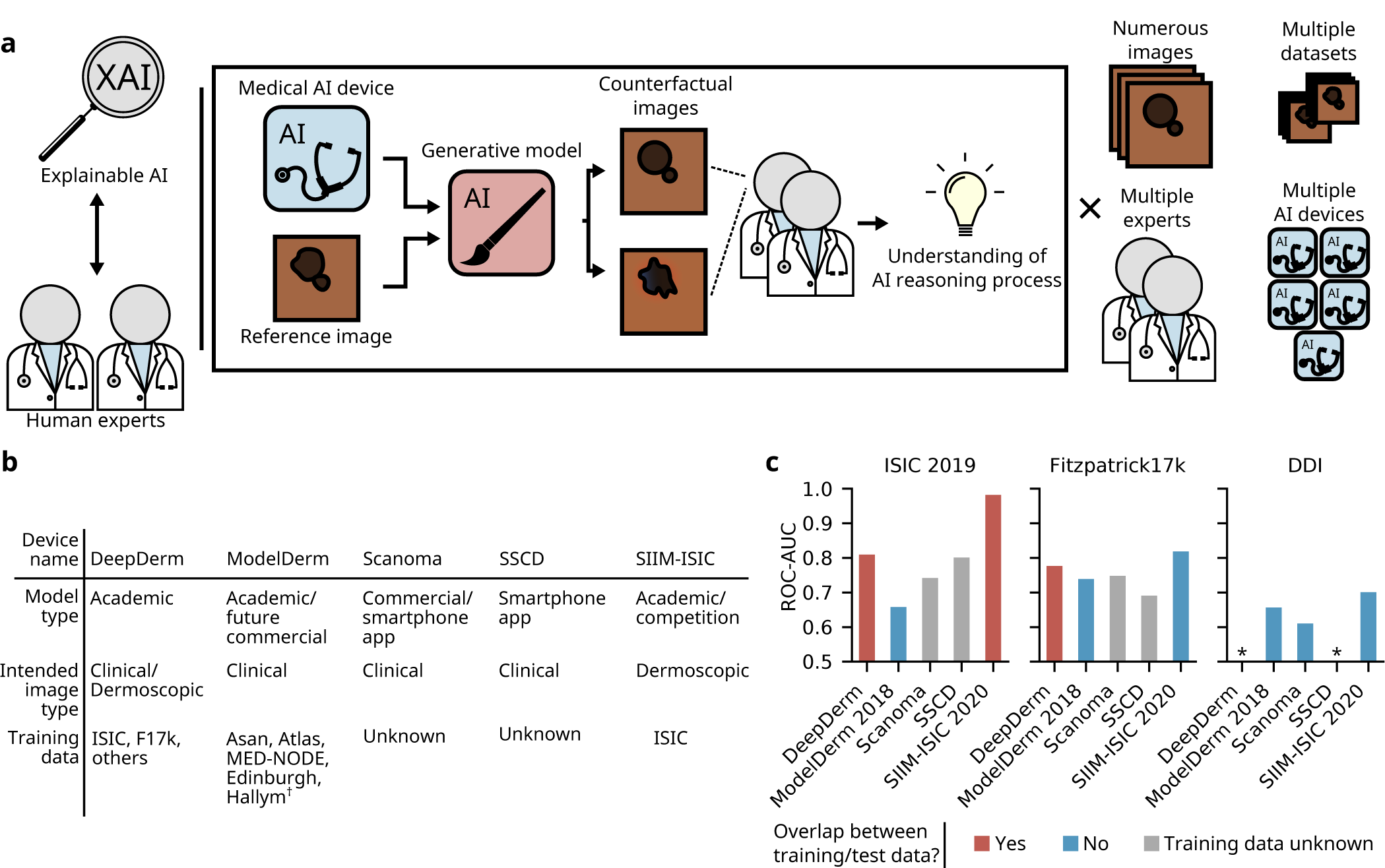
| Overview of joint expert, XAI auditing procedure and audited AI devices. **a,** Our auditing procedure unites explainable AI with analysis by human experts to understand medical AI devices. Specifically, we leverage generative models to create *counterfactual* images that alter the prediction a medical AI device; analysis of the counterfactuals by human experts (dermatologists) reveals the medical AI device’s reasoning processes. We perform the analysis on numerous images from each of multiple datasets, gathering insights from two experts, for each of five different dermatology AI devices. **b,** Key details of dermatology AI devices audited in this study. **c,** Performance of the dermatology AI devices on three datasets, including a dataset (DDI) external to the training data of every device. We examine the area under the receiver operating characteristic curve (ROC-AUC) to focus on the model’s internal reasoning processes rather than emphasize the authors’ original choices of model calibration. ^†^ Asan, Atlas, and Hallym datasets described in ref.^22^; MED-NODE is described in ref.^23^; Edinburgh is available at https://licensing.edinburgh-innovations.ed.ac.uk/product/dermofit-image-library *ROC-AUC*<*0.5 (i.e., worse than random performance).

## Results

### Overview of dermatology AI device selection and reproduction

Aiming to best represent the current state-of-the-art in dermatology AI devices, we explored the scientific literature and commercial market, ultimately choosing five AI devices to audit (Fig. 1b). These devices span the spectrum of academic and commercial devices, and include devices already distributed for use by consumers. The five devices are: (i) DeepDerm, a previously developed reproduction^21^—using the original training data—of the classifier from a seminal academic publication^15^, which hailed the classifier for its “dermatologist-level” performance; (ii) ModelDerm 2018^22^, an academic classifier for which a later version (which we were unable to obtain) was CE approved for use in the European economic zone; (iii and iv) Scanoma and Smart Skin Cancer Detection (SSCD), two consumer-facing, smartphone apps; and (v) a “competition-style” classifier, designed to mimic the key design decisions of the winning model^24^ from the 2020 SIIM-ISIC Melanoma Classification Kaggle challenge^25^ while circumventing that model’s prohibitive computational burden. Authors of additional AI devices declined to make available their full models (i.e., model weights), preventing us from analyzing other high-profile devices^16, 17^.

Since these diverse AI devices were trained on highly varied training data, we hypothesize they may exhibit a wide range of internal reasoning processes, for instance focusing on varied dermatological features or spurious signals. The training data include both dermoscopic images (taken through a specialized dermatological tool that magnifies and enables visualization of deeper layers of the skin) and clinical images (acquired with a digital camera, without the use of a dermatoscope). Dermoscopic and clinical images feature unique profiles of potential signals for AI systems to learn: for instance, dermoscopic images better reveal a lesion’s fine details, such as pigmentation patterns, and exhibit unique artifacts, such as ruler markings and dark corner artifacts; clinical images likewise may provide more information on a lesion’s context (location, surrounding lesions), in addition to their own characteristic artifacts, such as presence of markings or patient clothing. Dermoscopic images from the ISIC database^25–27^ were used to train both DeepDerm and SIIM-ISIC, though the particular subsets of data used for each model differed. DeepDerm also included clinical images in its training set, gathered from numerous online sources. ModelDerm trained on only clinical images, including publicly available images as well as images that were never made publicly available. The training procedures for the smartphone app AI devices have not been published, but based on the wide public availability of dermatology image datasets, we speculate they could have trained at least in part on images from ISIC, Fitzpatrick17k^28^, or other sources. Beyond the variability introduced by differences in training data, additional variation between the models may also arise from their diverse architectures, preprocessing schemes, ensembling, and other computational differences.

All of these devices aim to differentiate benign from malignant skin lesions, while some focus on the narrower problem of differentiating melanoma, the most deadly form of skin cancer, from melanoma look-alikes, such as benign nevi (moles), seborrheic keratoses, and dermatofibromas. We frame our analysis through this narrower problem, which has historically received great attention within the AI community, and which models a well-defined clinical task. In particular, we construct our test data to contain only melanomas and melanoma look-alikes, such that AI devices trained to more generally differentiate benign from malignant lesions here effectively function as melanoma classifiers. Since some classifiers were designed to function on dermoscopic images, others on clinical images, and at least one (DeepDerm) both, we examine all classifiers in each context, using ISIC as our source of dermoscopic images, and Fitzpatrick17k for clinical images (note that, since we are most interested in what alterations cause images to appear more benign or malignant and not benchmarking AI performance, we do not expect our XAI analysis to be sensitive to overlap between the training and test data)^8^.

We carefully adapted each AI device for use with our XAI tools, such that all analyses could be performed in a uniform software environment, thus eliminating a potential source of variation. Wherever feasible (*i.e.*, with the exception of SIIM-ISIC), we used the original model weights, to ensure that the original reasoning processes for that AI device could not change. While we suspect that the reasoning process of SIIM-ISIC should closely match the original 2020 SIIM-ISIC Kaggle competition winning model–we use the same training data, training procedure, and test-time image augmentations/ensembling–we intend our audit of SIIM-ISIC to shed light on the influence of these common, performance-boosting techniques rather than to definitively comment on the reasoning process of that original model. We verified our adaptations against the original implementations and achieved close reproduction of the original results; only slight differences arose due to platform-dependent implementation differences in preprocessing or arithmetic (Supplementary Fig. 2).

### Dermatology AI devices perform unreliably

As a first step toward understanding dermatology AI devices, we evaluated the performance of each device for differentiation between melanoma and melanoma look-alikes, finding the performance variable and often low (Fig. 1c). After accounting for train/test overlap (we expect AI devices trained on a particular dataset to perform artificially well), we note the following: (i) ModelDerm, which was trained only on clinical images, performs worst on the dermoscopic images (ISIC), though train/test overlap may unfairly advantage the other AI devices; (ii) Despite training on no clinical images, SIIM-ISIC outperforms all other models on clinical images; (iii) All models fail to achieve satisfactory performance on DDI, the only one of our three datasets known not to overlap with the training data of any AI device. This performance gap could come from DDI’s inclusion of diverse skin tones and rare diseases, but may also be due to other out-of-distribution features^21^. Our performance evaluation suggests that the five dermatology AI devices may rely on different internal reasoning processes, since the pattern of performance gains or losses across the three datasets does not hold consistent among the AI devices.

### Counterfactual images reveal basis for AI decisions

To understand the reasoning processes of the AI devices, we examined each AI device via an XAI tool: generation of counterfactual images. Counterfactual images reveal the basis of an AI device’s decisions by altering attributes of a reference image so as to produce a similar image that elicits a different prediction from the AI device. For instance, consider the case that an AI device predicts a lesion is malignant, while a counterfactual predicted by the AI device to be benign differs in that it features lighter, more uniform pigmentation, and fewer brown spots on the background skin; provided that we ensure all differences in the counterfactual push the AI device’s predictions in the desired direction (more benign), we may infer that the classifier uses darker pigmentation and brown spots on the background skin as part of its reasoning process (Fig. 2a).

**Fig. 2.**
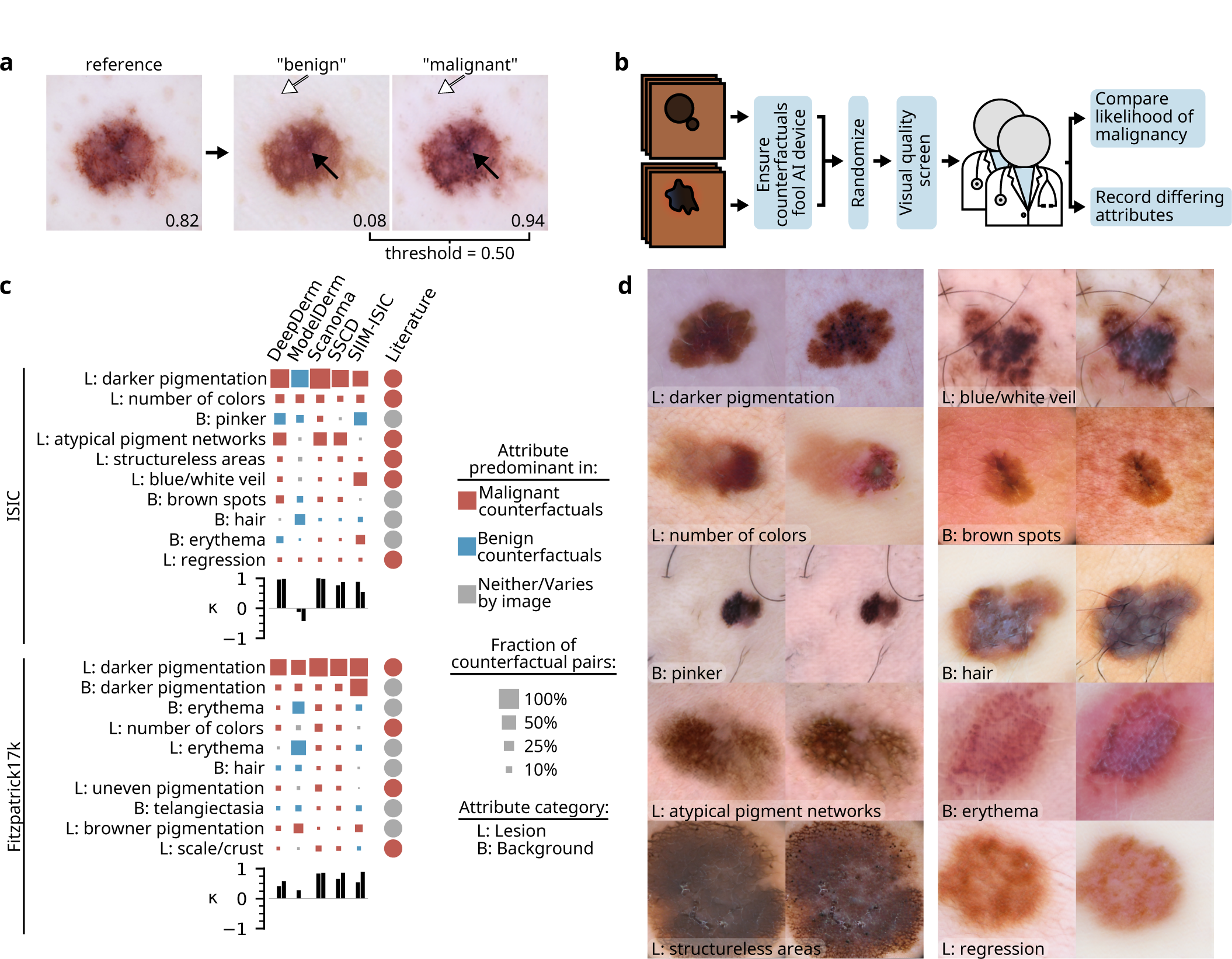
| Joint expert, XAI auditing procedure reveals reasoning processes of dermatology AI devices. **a,** Given a reference image and an AI device to investigate, our generative model produces “benign” and “malignant” counterfactuals, which resemble the reference image but differ in one or more attributes (e.g., pigmentation, solid arrows, and dots on the background skin, open arrows). When evaluated by the AI device, the counterfactuals’ outputs lie on opposite sides of the decision threshold. Higher values indicate greater likelihood of malignancy, as predicted by an AI device (Scanoma). **b,** To obtain robust conclusions, dermatology experts evaluate numerous counterfactuals after pre-screening and randomization of the images. **c,** Attributes identified by our joint expert-XAI auditing procedure as key influences on the output of dermatology AI devices. For each attribute/device pair, we count the proportion of counterfactual pairs in which experts noted that attribute differs; we display the global top-10 attributes as determined by lowest rank-sum over all AI devices. Based on expert evaluation of whether the attribute was present to a greater extent in the malignant or benign counterfactual of each pair, we determine whether that attribute was “predominant” in benign or malignant counterfactuals, i.e., present to greater extent in benign (malignant) counterfactuals in at least twice as many images as malignant (benign) counterfactuals. The size of each square is then determined as the number of counterfactual pairs with a difference noted in the predominant direction. For comparison, we specify how human dermatologists use each attribute (“Literature”), based on our review of the literature^29–35^ combined with expert opinion from two board-certified dermatologists; see Discussion for additional information. Bar charts indicate Cohen’s *κ* values for agreement between each expert and the AI device, where each is asked which image in each counterfactual pair appeared more likely to be malignant. “L”, lesion; “B”, background. **d,** Examples of counterfactuals that differ in each of the top ten attributes identified in the ISIC data; the attribute is present to a greater extent in the right image of each pair. For conciseness, some attribute names were shortened; refer to Supplementary Table 1 for full names. Images adapted with permission from ref.^27^ Combalia et al., ref.^26^ Tschandl et al., and ref.^36^ Codella et al.

To this end, we improved and applied a previously developed^8^ technique for generation of counterfactual images, Explanation by Progressive Exaggeration, with updates to enable more rigorous conclusions. In the context of our dermatology AI devices, this techniques enables generation of both “benign” and “malignant” counterfactuals from a reference image (Fig. 2a). We can then learn from comparing *two opposing counterfactuals*, which guards against potential misinterpretations, should the technique introduce any systematic changes to the counterfactuals. Explanation by Progressive Exaggeration trains a generative AI model in conjunction with an AI device, such that the generative model learns how to alter images to change the AI device’s predictions. We train the generative model to create counterfactuals that are similar to the reference image and appear realistic, but differ from the reference image in order to elicit the desired prediction from the AI device. Importantly, since the generated counterfactuals may alter more than one attribute, we updated the technique to ensure that we train the generative model to only change attributes when those changes elicit the desired effect on the AI device’s output, whereas the previously published version of this technique may also alter attributes irrelevant to the classifier’s output (Supplementary Fig. 3). Additional updates enabled generation of higher quality images that retain fine details, such as hair, that might be important for dermatology AI devices (Supplementary Fig. 4). We separately trained such generative models for each AI device, for each of the ISIC and Fitzpatrick17k datasets, for a total of ten generative models (Methods, Supplementary Fig. 5-6); a uniform set of training parameters facilitates comparison between the AI devices (Supplementary Fig. 7).

While examination of a single counterfactual pair provides some information about an AI device’s reasoning process, to obtain a more complete and rigorous understanding of the AI devices and enable direct comparisons between devices, we systematically interrogated thousands of counterfactual images, in a randomized and blinded fashion (Fig. 2b). We began our analysis by pre-screening the counterfactuals, to ensure we only examined high-quality counterfactuals and to facilitate comparisons between AI devices. We excluded counterfactuals that failed to produce the desired output from our AI devices (i.e., we ensured the “malignant” and “benign” counterfactuals lie on the correct sides of the decision threshold), or that contained visual artifacts (e.g., “water-droplet–like” artifacts^37^), as judged by dermatologists. Two dermatologists then independently annotated each counterfactual pair, which was randomized and blinded to reduce bias. To learn whether the dermatologists’ general impressions of the counterfactuals agreed with each AI device regarding what appears more or less malignant, we first inquired, “Which image appears most likely to represent a melanoma?” We then asked the dermatologists to record individual image attributes that differ between the “benign” and “malignant” counterfactuals, such that we could learn which attributes each AI device uses, and how it uses them (Supplementary Fig. 8).

We aggregated the dermatologists’ insights over thousands of counterfactuals to determine the reasoning process of each dermatology AI device. We conceptualize the reasoning process as swayed toward a benign or malignant prediction by key attributes identified as differing in counterfactual pairs; our analysis provides the typical direction of an attribute’s effect, based on whether that attribute was predominant in the benign or malignant counterfactuals, as well as an approximate idea of the extent of the effect, based on the frequency with which dermatologists observed that attribute differing in counterfactuals. Note that we expect this frequency to depend on multiple factors, including the fraction of the dataset to which that attribute is relevant, inductive biases of our generative models, and perhaps a combination of a dermatology AI system’s sensitivity to an attribute and the sensitivity of our evaluators in detecting that attribute (which may be at odds, in the case of a visually subtle change that sizeably affects a prediction). Our analysis reveals that the AI devices focus on both medically relevant and putatively spurious attributes, and exhibit considerable heterogeneity in how they interpret those attributes (Fig. 2c).

### A detailed view of medical AI reasoning

Our counterfactual analysis highlights the pigmentation of lesions as a key attribute in determining the predictions of all dermatology AI devices examined, for both dermoscopic and clinical images. In all cases, “darker pigmentation” surpassed all other attributes in frequency, with dermatologists noting this change in the majority of counterfactual pairs. Consistent with dermatologists’ interpretation of more darkly pigmented lesions, dermatology AI devices typically interpret darker pigmentation of lesions as increased likelihood of melanoma; the only exception is ModelDerm when evaluated on dermoscopic images—an image type upon which this model was never trained. A subset of the dermatology AI devices (DeepDerm, Scanoma, and SSCD) also base their decisions in part on atypical pigment networks for dermoscopic images, in agreement with dermatologists’ use of this attribute during pattern analysis of melanocytic lesions^29, 30^.

Dermatology AI devices also depend on a variety of other attributes of the lesion, many of which dermatologists also consider when analyzing melanocytic lesions. In both dermoscopic and clinical images, the AI devices consider the number of colors in a lesion, where a greater number of colors typically associates with predictions of malignancy^31^. Some AI devices, most prominently SIIM-ISIC, also consider the presence of a blue/white veil, which has previously been reported as a specific finding for melanoma^32, 33^. Other attributes of the lesion that factor into the AI devices’ decisions include presence of structureless areas or regression in dermoscopic images, and uneven pigmentation or erythema in clinical images. Aside from erythema, which varies between a benign or malignant signal depending on the AI device, these attributes typically associate with the malignant counterfactuals. Their frequency, however, varies considerably between devices, pointing out heterogeneity in the devices’ reasoning processes.

Analysis of each AI devices’s top attributes (Supplementary Fig. 9-10) revealed additional lesional attributes considered distinctively by only a subset of the AI devices. In dermoscopic images, these attributes included patchiness (DeepDerm and SSCD), strawberry pattern (ModelDerm), white spots (SSCD), prominence of follicles or pores (SSCD), white striae (SIIM-ISIC), and scale (SIIM-ISIC). In clinical images, these attributes included erosion or ulceration (DeepDerm and Scanoma), nodular or papular appearance (ModelDerm), uneven borders (ModelDerm), and the shininess of a lesion (SIIM-ISIC).

Attributes of the background skin also influence the dermatology AI devices, and in comparison to attributes of the lesion, often elicit more diverse responses among the devices: Brown spots on the background skin influence towards benign or malignant predictions depending on the device. Hair typically associates with benign counterfactuals in dermoscopic images, but can also associate with malignant counterfactuals in clinical images. More textured skin (e.g, skin grooves) associates with the benign counterfactuals of Scanoma and ModelDerm (Supplementary Fig. 9), but is rarely highlighted by the counterfactuals of other devices. Erythema or telangiectasias of the background skin also feature prominently in the results of our counterfactual analysis, and the effects of these attributes vary both between AI devices and within an AI device, depending on whether an image is clinical or dermoscopic. Finally, counterfactuals highlighted the “pinkness” of background skin as influencing AI devices’ decisions, particularly in dermoscopic images. In contrast to erythema, this attribute often applies uniformly across an image (Fig. 2d), consistent with effects of lighting or an image’s color balance. Similarly, we recorded overall darker images and cooler color temperatures as influential for one classifier (SIIM-ISIC). Similar to other background skin attributes, lighting or color balance changes may sway an AI device toward a more benign or more malignant prediction depending on the device. In comparison, we were unable to identify dermatological literature that establishes these attributes of the background skin as signals commonly used by dermatologists.

Darker pigmentation of the background skin, which stands out as the overall second most frequently recorded difference in our clinical counterfactuals, consistently associates with malignant counterfactuals. We observed that the darker pigmentation sometimes localized to discrete areas of the background skin, for instance to the immediate periphery of a lesion (effectively enlarging the lesion), or alternatively to areas of the image in shadow. In other instances, darker pigmentation extended more uniformly throughout the background skin. Among the classifiers, SIIM-ISIC—a model that was trained on only images of light skin, and the only of our devices known not to include clinical images in its training data—featured this attribute most prominently in its counterfactuals, though all classifiers were sensitive to this attribute.

In general, AI devices and human dermatologists agreed on which image in the counterfactual pair most likely de-picted a malignancy. The exception, ModelDerm, exhibited negative Cohen Kappa values compared to dermatologists on dermoscopic images; its interpretation of key attributes, such as darker pigmentation of the lesion, diverged from the other devices. This device also agreed poorly on clinical images, again coinciding with its focus on a unique profile of attributes. Curiously, Scanoma achieved the best agreement with dermatologists on both datasets, despite other AI devices achieving higher predictive performance (even when that performance was on external data and therefore not inflated by train-test overlap, e.g., SIIM-ISIC with Fitzpatrick17k; Fig. 1c).

### Validation of insights from counterfactuals

While we engineered our counterfactual generation procedure to ensure that detected attributes indeed influence AI devices’ predictions, we performed additional analyses to verify these conclusions. Ideally, we may confirm our findings by performing a targeted intervention to experimentally modify a single attribute of an image, in a well-defined fashion, then monitor the intervention’s effect on each AI device’s prediction. While existing techniques do not enable reliable modification of most attributes detected in our analysis (e.g., addition or removal of atypical pigment networks without altering other attributes), well-established techniques enable programmatic modification of the color of an image, enabling us to experimentally produce images that are more or less “pink”, an attribute detected as influential to most classifiers (Fig. 2c and Fig. 3a). We shifted the color (i.e., the u’ and v’ chromaticity coordinates in the CIELUV color space^38^) of each image in the ISIC dataset, then monitored how each AI device’s prediction changed for a range of colors (Fig. 3b).

**Fig. 3.**
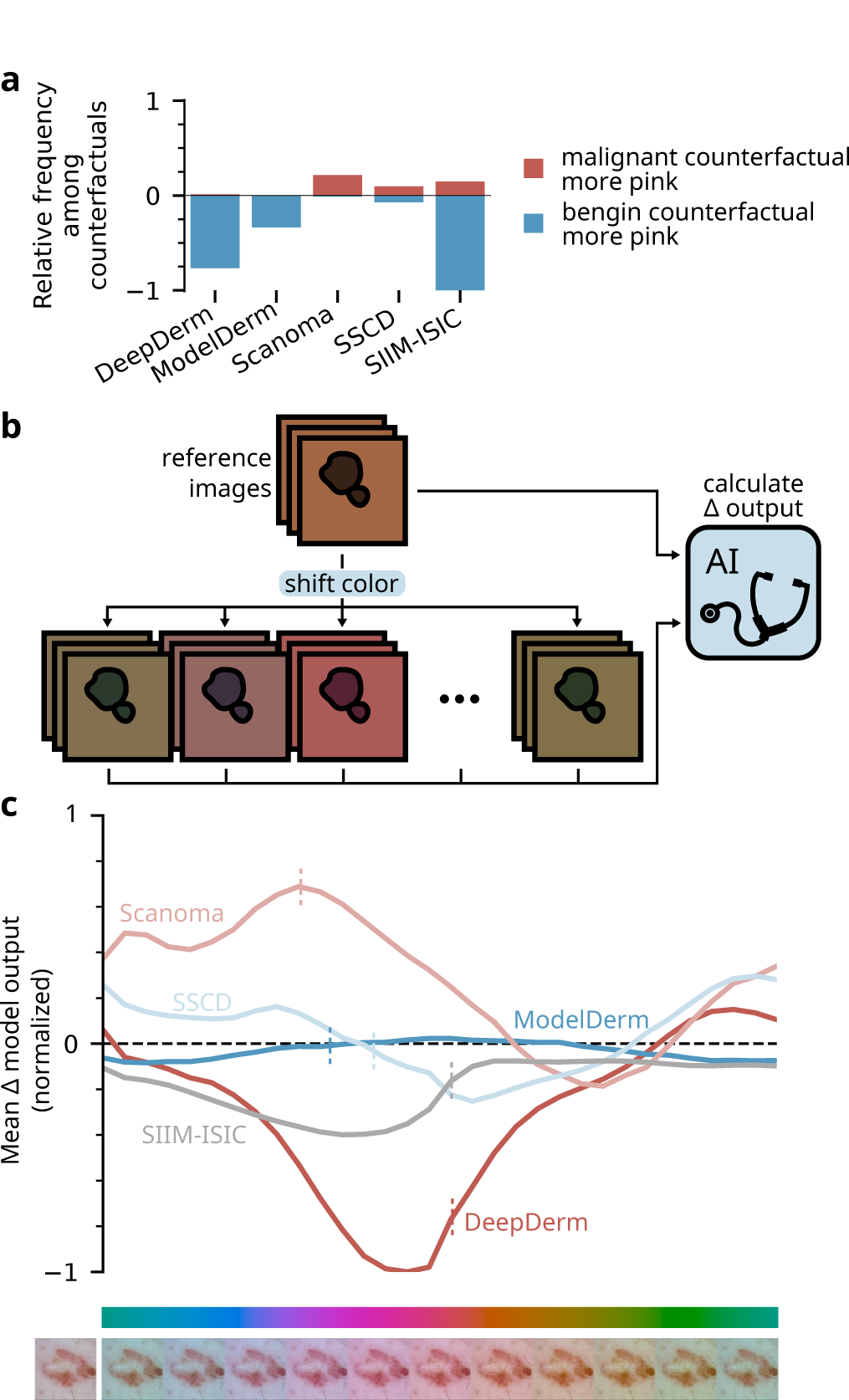
| Experimental validation of findings from expert analysis of counterfactual images. **a,** Frequency with which experts noted that either the benign or malignant image in a pair of counterfactuals displayed a pinker background; this view details our observations from the ISIC dataset summarized in Fig. 2c, in the row “B: pinker”. The vertical axis is normalized relative to the maximum observed frequency, that is, 42% of counterfactual pairs from SIIM-ISIC. **b,** Experimental setup used to verify the importance of a pink tint to the AI devices’ predictions. We programmatically color-shifted each image in the ISIC dataset (*n* = 20260) by modifying its chromaticity coordinates in the CIELUV color space (see Methods), then compared each AI device’s predictions between the original and color-shifted images. **c,** Sensitivity of each AI device to programmatic color shifts, mirroring observations from our counterfactual experiments regarding the effect of pinker tints on the AI devices’ predictions. The vertical axis is normalized relative to the maximum change in AI device output, *i.e.*, a decrease of 0.17 with DeepDerm. Vertical dashed lines indicate the mean change in chromaticity (color) among counterfactual pairs annotated as differing in their pink tone. Example color-shifted images (below color bar) display the extent of the color shift; the reference image, adapted with permission from the ISIC archive^36^, appears at far left.

These experimental modifications of image color and their impact on the predictions of the AI devices recapitulates the trend observed in our previous analysis of counterfactual images (Fig. 3c; compare to 3a): e.g., pinker images elicit more benign predictions from DeepDerm and more malignant predictions from Scanoma. Multiple factors including the “sensitivity” of an AI device to changes in an attribute determine the relative frequency of an attribute among counterfactuals (Fig. 3a); thus, magnitudes are not directly comparable (see Results: “Counterfactual images reveal basis for AI decisions”). This experiment validates that the attributes identified in our previous analysis of counterfactual images indeed influence the output of the AI devices in the direction described by the counterfactual analysis. In addition, this experiment validates our interpretation of “pinker background skin” as a global change in lighting or color balance. Indeed, our experimental procedure mirrors computational techniques used to perform white balancing (correction for chromatic adaptation) in digital cameras and highlights how changes to lighting or camera settings might affect AI dermatology devices’ predictions in undesirable ways.

### Counterfactuals explain failure cases

To reinforce the core findings from our systematic analysis of counterfactuals, we also present counterfactual explana-tions of cases in which the AI devices failed to correctly predict whether a lesion was malignant or benign.

The reliance of dermatology AI models on the pigmentation of a lesion can lead to failures that are “reasonable”, in that they might also be expected from human dermatologists (Fig. 4a): for instance, while presence of atypical pigment networks and darker pigmentation lead one AI device to predict a lesion was malignant, it turned out to be benign; indeed, authors of this present study who practice dermatology find this lesion concerning for the same reason, and would have opted to biopsy the lesion.

**Fig. 4.**
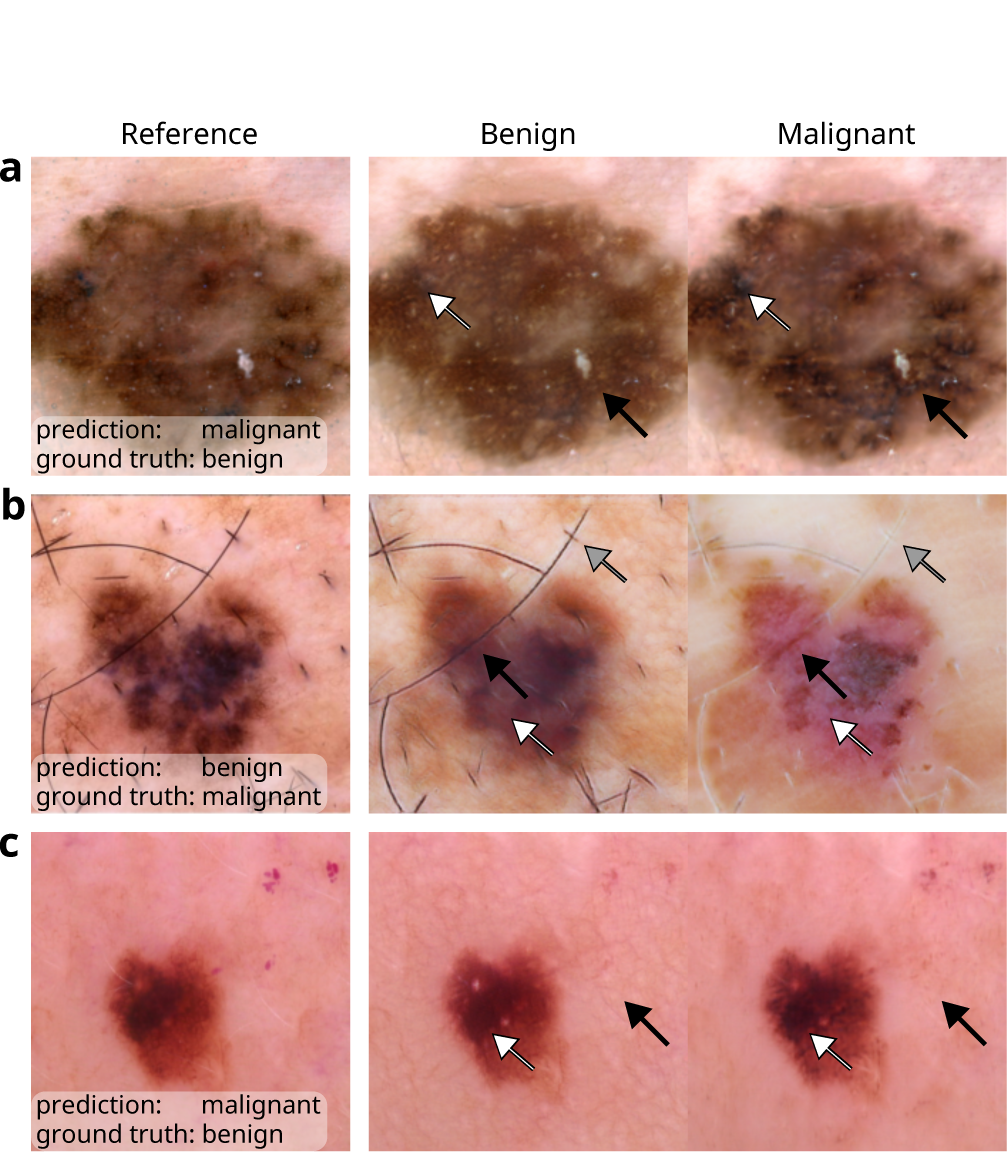
| Explanations of failure cases of dermatology AI devices, illustrating key findings from our systematic analysis. **a,** Presence of atypical pigment networks (black arrows) and darker pigmentation (white arrows) contributed to a false positive prediction from Scanoma. **b,** Curiously, ModelDerm may have required lighter pigmentation (black arrows), increased erythema (white arrows), and less hair on background skin (gray arrows) to correctly predict this image pictures a melanoma. **c**, Lack of prominent skin grooves or reticulation on the background skin (black arrows), alongside darker pigmentation (white arrows), contributed to another false positive prediction from Scanoma. Images adapted, with permission, from the ISIC archive^26, 27, 36^.

In other cases, dermatology AI models rely on potentially relevant attributes of an image, but use these attributes incorrectly. ModelDerm misclassified a malignant lesion as benign, and examination of the corresponding counter-factuals revealed attributes such as darker pigmentation of the lesion and absence of erythema as influential for this decision (Fig. 4b). However, dermatologists would not typically associate darker pigmentation with decreased likeli-hood of melanoma, and the increased erythema of the malignant counterfactual is more consistent with the “strawberry pattern” of facial actinic keratoses, a type of premalignant skin lesion^30^.

Dermatology AI devices also utilize likely irrelevant attributes in their reasoning process, including associating hair on background skin with benign lesions (Fig. 4b). In another example (Fig. 4c), a classifier misclassifies a benign lesion as melanoma in part due to the texture of the background skin, namely its lack of prominent skin grooves or reticulation.

## Discussion

Relative to previous techniques to analyze medical image AI devices, our framework provides numerous advantages, which together enable us to present the most detailed view to date of the reasoning processes of AI systems for medical images. Whereas the *de facto* standard XAI technique for image models, saliency maps, best reveals the importance of localizable attributes, our discovery of dependencies on numerous overlapping, textural, and tonal changes to an image showcases the importance of our use of XAI based on counterfactual images, and highlights limitations of previous work that relied only on saliency maps^5^. In fact, we surmise that most attributes identified by our framework, such as darker pigmentation of lesions, number of colors in a lesion, presence of erythema, pigmentation patterns, etc., would be unlikely to be identified by saliency maps. Our framework also improves upon previous efforts^8, 9^ to analyze medical image AI systems via counterfactual images. In contrast to other generative techniques^8, 9^ for counterfactual generation (including the original Explanation by Progressive Exaggeration) or simply comparing real images predicted as benign and malignant, our method enables the inference that each attribute that differs in a benign/malignant pair is indeed important for the AI device’s predictions (Supplementary Fig. 3). Our method also offers more detailed reproduction of fine-grained features such as hair (Supplementary Fig. 4), which we discovered to influence some AI devices. Perhaps more importantly, our framework introduces a means to translate XAI outputs to a human-understandable, medically meaningful form, namely via systematic, randomized, blinded analysis by medical experts. Particularly for a high-stakes application such as medical decision-making, we contend that such a medically-grounded understanding offers greatest potential for actionability.

We find that dermatology AI devices leverage a number of medically meaningful attributes found within lesions— including attributes related to a lesion’s pigmentation—in a manner consistent with human experts. Dermatology AI devices also rely on numerous attributes with debatable medical relevance and unclear desirability. Brown spots on the background skin may signify a patient’s age or history of sun exposure (a risk factor for melanoma^39^) but are not in any established melanoma diagnosis guidelines. Erythema, particularly in a “pink rim” distribution around a lesion^34^, has been associated with melanoma, but also with benign melanoma look-alikes such as irritated seborrheic keratoses^35^. Hair may suggest a lesion’s location on the body while skin grooves may provide clues on a lesion’s location (e.g., acral), the patient’s age, or history of sun exposure. Lighting conditions or color balance also influence many dermatology AI devices, and we surmise these almost certainly undesirable dependencies arise from spurious differences in image acquisition or preprocessing. Beyond the fundamental scientific interest of this detailed characterization of AI reasoning processes, our approach could be used by AI developers to improve their models and to inform stakeholders on the trustworthiness of medical AI devices.

This methodology can help uncover idiosyncratic failure modes of AI, with implications for its regulation and medical use. We expect distributional shifts in medical AI to be common–especially in dermatology AI, given the diversity of image acquisition devices, lighting conditions, skin appearances across demographics, and lack of implemented image standards. Our findings suggest that common distributional shifts, such as changes in lighting or color balance, will alter AI performance. Thus, we caution potential users of such devices that a device’s advertised performance, which is often estimated in a well-circumscribed setting, may not be achieved in real-world use^21^. Our findings also imply that regulators should scrutinize the distribution of data on which a device is evaluated, with particular attention toward (i) ensuring it well reflects the intended deployment distribution, and (ii) considering differential performance across subgroups (e.g., varied acquisition devices or regions, or key potential dependencies such as lighting and skin tone). For AI developers, we envision that our methodology may enable more tractable debugging of AI devices prior to more expensive and time-consuming multi-site performance evaluations^40^.

A previous publication highlighted that dermatology AI devices perform worse on darker skin tones^21^, and our study reveals *mechanistic* insights on how this bias may arise in the reasoning processes of dermatology AI devices. Moreover, contrasting with that study’s focus on a single source of potential bias (skin tone), our method uncovers this bias in an untargeted manner. With our methodology, evaluators often noted diffusely darker background skin in the malignant counterfactuals (especially those of SIIM-ISIC, which was trained on only images of light skin). The real-world variations most likely to produce changes similar to those observed in the counterfactuals include lighting conditions, camera settings (e.g., exposure and color balance), and variations in skin tone. Since the generative models may entangle attributes that arise from different physical origins (i.e., skin tone and lighting), the counterfactuals do not enable us to distinguish between these (non-mutually exclusive) possibilities, but both are concerning. First, to the extent that real-world variations in skin tone may recapitulate the darker background skin observed in our counterfactuals, dermatology AI devices may exhibit a direct dependence on skin tone, where darker skin elicits more malignant predictions. Second, even if AI devices do not depend directly on skin tone, sensitivity to lighting conditions or camera settings may also introduce an indirect dependence on skin tone: camera designs are often biased toward ensuring appropriate color in light skin tones, but not dark skin tones^41^, implying that the dependence of AI devices on lighting or color balance may manifest systematically in images of dark skin. Similarly, our counterfactuals occasionally highlighted reflections as influential, which could systematically bias predictions in images of dark skin acquired with suboptimal lighting (e.g., use of camera flash)^42^. These findings reinforce that developers should ensure that dermatology AI performs well on dark skin, which is often under-represented in dermatology databases^43^, and highlight the importance of high quality, alongside quantity.

In conceptualizing the reasoning processes of dermatology AI devices, we aimed to characterize the devices in medically meaningful, human-derived terms, while retaining flexibility to faithfully represent processes that *a priori* need not coincide with human concepts. We therefore did not limit our set of attributes to any predefined list and instead enabled our evaluators to input any attribute they noticed and could describe. A limitation of this approach is that human biases may nonetheless prevent our analysis from uncovering peculiar, AI-specific patterns. Furthermore, our analysis does not attempt to provide a complete picture of the decision boundary of the AI devices. We instead characterize their reasoning processes with respect to a particular distribution of images, namely, realistic dermoscopic and clinical dermatological images, implying that our analysis thus provides limited information on out-of-distribution images, or features that rarely appear in the examined images (e.g., patches) . However, our choices to frame our analysis in terms of (i) human-derived concepts and (ii) a distribution of images that approximates a clinical use case, enable more medically meaningful inferences on the AI devices’ reasoning processes and how they could lead to desirable or undesirable behavior in deployment.

In addition to the immediate value of our analysis to understanding dermatology AI devices, our analysis provides a general framework for auditing complex AI systems that require specialized domain knowledge to best understand. Specifically, investigators could apply our complete analysis pipeline—training of generative models to synthesize counterfactuals, querying of experts via a randomized and blinded data collection app using freeform “attribute” fields, and compilation of those responses to attain a global understanding of an AI system—to characterize other AI medical image analysis tools, such as the numerous AI-based medical image analysis systems that have been deployed clinically, as well as for non-medical, computer-vision tasks such as facial recognition, scene classification in autonomous vehicles, or industrial or agricultural monitoring. In addition, our framework for querying experts and compiling responses could be applied in conjunction with other XAI techniques to understand AI systems outside the image domain, in cases where input features still lack stable semantics, such as systems that operate on time-series data. More generally, our study sets a precedent for rigorous application of explainable AI, addressing key issues that may have imperiled previous XAI analyses: insufficient sampling, potential for bias, lack of expert involvement, and failure to examine AI systems in multiple contexts.

## Methods

### Image selection and preprocessing

To interrogate the performance of AI-based dermatological classifiers, we collected images of melanomas and melanoma look-alike lesions from multiple sources. Our first source, Fitzpatrick17k^28^, consists of clinical (rather than dermoscopic) images previously aggregated from online dermatology atlases. We filtered Fitzpatrick17k to include only melanomas, benign melanocytic lesions, seborrheic keratoses, and dermatofibromas. We additionally excluded dia-gramatic and histopathological images, and images that could be clearly identified as pediatric; after exclusions, the dataset consisted of 889 images. Advantages of Fitzpatrick17k include closer approximation of the expected inputs to consumer-facing dermatology AI tools (as compared to dermoscopic images, which require specialized tools) and inclusion of a variety of skin tones. Disadvantages include its relatively small size after filtering and noise in the diagnosis labels, which may not have been acquired via histopathological analysis or other gold-standard means.

Our second source, the ISIC 2019 challenge dataset^26, 27, 36^, consists of dermoscopic images from a variety of primary sources, including HAM10000^26^ and BCN20000^27^. Like Fitzpatrick17k, we filtered the dataset to include melanomas, as well as melanoma look-alikes: benign melanocytic lesions, seborrheic keratoses, and dermatofibromas. After filtering, the ISIC dataset consisted of 20260 images. Most lesions were confirmed via histopathology (n=13072) or serial imaging showing no change (n=3704), while a smaller number were confirmed by single image expert consensus (n=1207), confocal microscopy with consensus dermoscopy (n=712), or unspecified means (n=1565). Compared to Fitzpatrick17k, ISIC thus offers more reliable diagnoses, but it lacks diversity in skin tones, featuring predominately light skin.

Finally, our third source, DDI^21^, consists of clinical images gathered from Stanford Clinics. Like other datasets, we filtered DDI to include only melanomas and melanoma look-alikes. In the case of DDI, which contains more granular and varied diagnoses, we included the following labels in our “melanoma” category: acral lentiginous melanoma, melanoma *it situ*, nodular melanoma, as well as the general tag “melanoma”. As melanoma look-alikes, we included the following labels: acral melanotic macule, atypical spindle cell nevus of reed, benign keratosis, blue nevus, dermatofibroma, dysplastic nevus, epidermal nevus, hyperpigmentation, keloid, inverted follicular keratosis, melanocytic nevi, nevus lipomatosus superficialis, pigmented spindle cell nevus of reed, seborrheic keratosis, irritated seborrheic keratosis, and solar lentigo. After filtering, DDI included 282 images; due to the comparatively high volume of data required for training our generative models, DDI was used only for performance evaluation (Fig. 1), rather than for our in-depth analysis of medical AI reasoning processes. However, DDI offers a number of desirable characteristics for evaluation purposes: (i) its images were not publicly available until after we obtained the five audited dermatology AI devices, precluding train-test overlap; (ii) DDI images have diverse skin tones, including enrichment for Fitzpatrick skin types V and VI; (iii) DDI contains a wide variety of skin conditions, including uncommon conditions; and (iv) the lesions are histopathologically proven, guaranteeing label accuracy. We note also that DDI is likely enriched for challenging lesions, since these are the lesions likely to require a biopsy.

### Classifier reproduction

We reproduced five AI-based dermatological classifiers, including prominent academically designed classifiers proposed for clinical use and classifiers currently in use by the public. Two of the classifiers, *Scanoma* and *Smart Skin Cancer Detection* (SSCD) are designed for use on mobile devices by the general public. The DeepDerm classifier is a previously published reproduction^21^ of a prominent academic model^15^, sharing its training data and architecture. The ModelDerm 2018 classifier is a publicly distributed academic model^22^, of which a later iteration (for which model weights are not publicly available) has been CE marked for use by the general public in Europe. The SIIM-ISIC Kaggle competition classifier is a reproduction of the first-place classifier^24^ in the 2020 SIIM-ISIC Kaggle competition^25^. These models cover a broad range of architectures, pre-processing techniques, and training data sources; as such we believe these models offer a thorough view of both current practices and the state-of-the-art in dermatology AI.

Scanoma is commercial software available for mobile platforms including iOS and Android; at the time of writing, the app’s AI classifier is free to use, while follow-up human evaluation is available for a fee. Architecturally, it is a custom convolutional neural network consistent with a MnasNet^44^, that is further optimized for use on mobile devices via quantization^45^. We obtained and unzipped the Scanoma APK file (normally installed on Android devices) to examine its TensorFlow Lite (TFlite) file, which contains the model specification and weights. Since our analysis tools are based on the PyTorch software library, we converted the network to the cross-library Open Neural Network Exchange (ONNX) format, which we then parsed in PyTorch. To maintain consistency with the original, quantized network while maintaining useful gradients, we implement the network using “fake quantization”^45^. We verified that our PyTorch re-implementation matches the TensorFlow Lite implementation by comparing a series of 1000 test images, and we achieved nearly identical outputs (r=0.99, Supplementary Fig. 2a). To account for the small discrepancy between the classifiers, we analyzed the processing pipeline step-by-step and found slight differences in the bilinear rescaling preprocessing step, which may differ due to different antialiasing constants; the remaining differences were explained by sporadic single-bit differences in the quantized feature maps, likely resulting from numerical differences between TensorFlow Lite’s native integer arithmetic routines and the equivalent operations performed in floating point arithmetic followed by fake quantization.

Like Scanoma, SSCD is a publicly available app intended for use on mobile devices. The architecture is a MobileNetV1, evaluated using floating-point (non-quantized) arithmetic. We followed a similar process to re-implement the SSCD classifier in PyTorch: a TFLite file was obtained from the app’s APK package, then converted to ONNX before loading in PyTorch. We again verified our reproduction using a series of 1000 images and found that our PyTorch re-implementation of the neural network exactly matched the original Tensorflow Lite network. However, to ease comparison between classifiers, we update the input image resizing routine (a pre-processing step, prior to the neural network) in our implementation relative to the original app. Whereas the original app asks a user to specify a bounding box and then scales this box to the 224*×*224-pixel input image (warping the aspect ratio), we use the same preprocessing routine as for all other networks, in which we first center-crop the image and then resize the image using a bilinear filter. To assess the impact of this change in image preprocessing, we compared our PyTorch implementation against (i) the original TFLite model accompanied by preprocessing with square center-cropping and nearest-neighbor resizing and (ii) the original TFLite model with variable aspect-ratio resizing using nearest-neighbor rescaling (matching the original Android implementation, under the assumption that the uncropped image represents a user-defined bounding box), and we observed Pearson correlation coefficients of 0.97 and 0.92, respectively (Supplementary Figs. 2b-c). While evaluation of the entire processing pipeline including user selection of bounding boxes and choice of resampling filters is important for clinical evaluation of an AI system, our study instead focuses on the decision-making processes of the neural networks.

ModelDerm^22^ is an academic classifier that has undergone multiple iterations, some of which have been tested in clinical settings, and one version of which has been approved for use in Europe via CE marking. We analyze the latest version for which model weights are publicly available, which we term ModelDerm 2018 based on the date of the accompanying publication^22^; authors declined to provide weights for the latest version of the model due to commercialization plans. ModelDerm is a ResNet-152^46^ that runs natively in PyCaffe, with preprocessing performed in OpenCV. We parse the model architecture and weights directly from Caffe Protocol Buffer files and reconstruct the model in PyTorch. While the majority of the processing pipeline is highly reproducible in PyTorch relative to the original implementation, the original implementation preprocesses images channel-by-channel using the histogram equalization function in OpenCV, which we could not exactly reproduce in PyTorch while maintaining meaningful gradients during backpropagation. Instead, we implemented a custom, differentiable analogue of histogram equalization, in which the empirical cumulative density function used in OpenCV’s implementation is replaced with a piecewise-linear approximation. Our PyTorch reimplementation of ModelDerm 2018, including the differentiable histogram equalization preprocessing step, retains close correspondence to the original PyCaffe/OpenCV implemention (r=0.96, Supplementary Fig. 2d).

The SIIM-ISIC competition classifier is intended to represent key features responsible for the high performance of the first-place winning classifier from the 2020 SIIM-ISIC melanoma classification Kaggle challenge, while reducing the computational complexity to permit feasible analysis. The original classifier is an extremely large ensemble of 90 networks, comprising mostly EfficientNets^47^, but also a few SE-ResNext 101s^48^ and ResNest101s^49^, all of which are evaluated at test time on 8 flips and rotations of the test image, for a total of 720 model evaluations per prediction. We reduced the computational complexity by retraining an ensemble of 3 EfficientNets (an EfficientNet-B5, -B6, and -B7), which comprise 80 of the 90 classifiers in the original ensemble, using the same training data, augmentation scheme and hyperparameters as the original classifiers. Our classifier additionally retains 8-fold image augmentation at test time, which we suspected may reduce the classifier’s sensitivity to subtle image variations. While not intended to be an exact reproduction of the original winning classifier, our classifiers attain only slightly lower classification performance in 5-fold cross validation as compared to the original classifier (area under the receiver operating characteristic curve of 0.966 vs. 0.985).

The DeepDerm classifier is a previously published reproduction^21^ of an academically developed model that was acclaimed for performing similarly well to dermatologists^15^. DeepDerm shares the same architecture (Inception-V3^50^) and importantly, the same training data as the original model, which was not publicly released. Since DeepDerm is distributed natively in PyTorch, no conversion steps were necessary for this classifier.

### Counterfactual generation

To identify specific image factors responsible for each classifier’s predictions, we generated counterfactual images using a variant of the technique “Explanation by Progressive Exaggeration”^8^. However, to improve image quality, stabilize training, and better restrict generated alterations to those that cause a classifier to output a different prediction, we introduce multiple updates. We begin with an overview of the technique, then explain our specific updates.

Explanation by progressive exaggeration uses generative adversarial networks to create alternate versions of images that (i) appear “realistic”, in the sense that they lie on the manifold of training images, (ii) produce the desired target prediction from a classifier, such as a prediction on the opposite side of the decision threshold as the original image, and (iii) are similar to the original image, in the sense that the original image may be approximately reconstructed by passing an altered, generated image back through the generator.

Formally, let *X* ⊂ [0, 1]^*d*^2^^ represent a set of images drawn from some data manifold *M*_*x*_, where *d* ∈ ℕ is the horizontal and vertical resolution of the (square) images, and let *f* : [0, 1]^*d*^2^^ *→* [0, 1] be a classifier to be audited. Our goal is to obtain a generator *G* : [0, 1]^*d*^2^^ *× C →* [0, 1]^*d*^2^^ that produces a counterfactual image 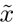 when given an input image *x* and a condition *c ∈ C* ⊂ ℕ, which indicates the target output that the classifier should produce when evaluated on the counterfactual image 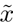. (Note that for simplicity of notation, we condense the generator and encoder of the original paper into a single function *G*). As in the original implementation of explanation by progressive exaggeration, our condition *c* is a discrete value that indexes a “bin” in the discretized output space of the classifier *f* ; we chose *C* = {0, 1*, …,* 9} with corresponding target outputs in the bins {[0, 0.1), [0.1, 0.2)*, …,* [0.9, 1]}. The three requirements listed above then translate to (i) the range of the generator *G*(*X, C*) is contained in the data manifold *M_X_*, (ii) the prediction of the classifier for the generated image *f* (*G*(*x, c*)) is approximately equal to the target output (in our case, the bin’s center at *c/*10 + 0.05), and (iii) if *f* (*x*) falls within the bin indexed by *c*, then *G*(*G*(*x, c^′^*)*, c*) *≈ x* for each *c^′^ ∈ C*.

To obtain a generator with these properties, we optimize the generator *G* in conjunction with a discriminator network *D* : [0, 1]*^d^*^2^ *→* ℝ that attempts to distinguish real from generated images. In contrast to the original implementation, we update the discriminator such that it does not depend on a condition *c*. The original implementation of the discriminator attempts to differentiate generated images from real images that elicit a particular prediction from the classifier, which may encourage generated images to appear similar to that subset of real images including potentially via changes that do not alter the output of the classifier. In contrast our implementation of the discriminator instead attempts to differentiate generated images from any real image, such that it only encourages that the generated images appear similar to real images (Supplementary Fig. 3). To reflect this update, we choose the following functions for the loss of the discriminator *L_D_* and of the generator *L_G_*. In the following equations, the random variables *X* and *C* take values in *X* and *C* and are distributed uniformly over *X* and *C*; *θ_D_* and *θ_G_* are the parameters of the discriminator and generator, respectively; *b* : [0, 1] *→ C* returns the bin index *b*(*f* (*X*)) of the output of the classifier; 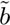 : *C →* {0.05, 0.15, …, 0.95} returns the center of the bin at index *C*; and *D_KL_* is the Kullback–Leibler divergence:

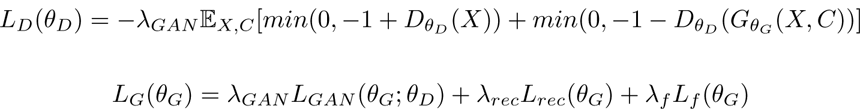

The individual components of *L_G_* are as follows:

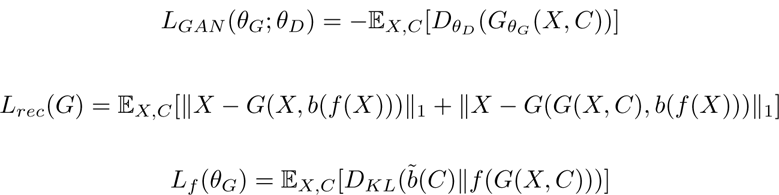

In addition our introduction of a non-conditional discriminator, we also update *G* to use an architecture similar to that used in CycleGANs^51^. This network is similar to the residual network-based autoencoder used in the original implementation of explanation by progressive exaggeration, but we found it produced images of higher visual quality (Supplementary Fig. 4).

To optimize the networks, we followed the reference implementation and used an Adam optimizer with a learning rate of 2 *×* 10*^−^*^4^, *β*_1_ = 0, and *β*_2_ = 0.9, with a mini-batch size of 32. To prevent the discriminator from outpacing the generator, we trained the discriminator for 5 mini-batches for each mini-batch that the generator was trained, and we applied spectral normalization to the discriminator’s parameters. To prevent overfitting, we also applied data augmentation including random cropping and random brightness modifications. To choose the hyperparameters *λ*, we followed the original publication and chose *λ_GAN_* = 1 and *λ_f_* = 1. To balance the magnitude of the generator’s alterations such that the counterfactuals were similar to original images but still contained perceptible differences (based on manual visual analysis of images), we chose *λ_cyc_* = 3 after gradually relaxing the *λ_cyc_* term from the value *λ_cyc_* = 100 suggested in the original publication (Supplementary Fig. 7). The generative models for each classifier and for each dataset were all trained using identical parameters. Comparison of counterfactuals generated by independent re-trainings of a generative model preserved which attributes varied between the benign and malignant counterfactuals (Supplementary Fig. 11), so we focused on a single generative model for each combination of AI device and generative model (Supplementary Table 2).

To train our models, we reimplemented the original TensorFlow library for explanation by progressive exaggeration using PyTorch. Generative models were trained for either 500 epochs (ISIC dataset) or 10^4^ epochs (Fitzpatrick17k dataset), to achieve approximately equal total training time for each dataset (*∼*10,000 kilo images); training time for a single generative model amounted to between one week and one month on an NVIDIA RTX 2080 TI graphics processing unit, depending on the complexity of the classifier.

### Expert evaluation of counterfactuals

To identify specific image factors upon which dermatological classifiers base their predictions, we asked two board-certified dermatologists, each with six years of experience, to analyze generated counterfactual images and determine which aspects of each image were altered, implying that they affect the classifiers’ decisions. We queried these dermatologists on hundreds of pairs of counterfactuals for each of five classifiers and two image datasets, amounting to thousands of responses. Each pair of counterfactuals was generated from a common “reference” image and consisted of an image that the classifier predicted to appear more benign, and an image that the classifier predicted to appear more malignant, such that both images depicted the same lesion but displayed differences that altered the output of a classifier.

To facilitate interpretation of the dermatologists’ responses and comparison of the classifiers, we prescreened the counterfactual images before analysis of the alterations within counterfactual pairs. Our prescreening consisted of a “classifier-consistency” criterion to ensure that the alterations between each pair of counterfactuals meaningfully changed the classifiers’ predictions, and a “visual quality” criterion to mitigate the presence of artifacts, which could impede our ability to infer the importance of non-artifactual alterations. Our classifier-consistency criterion required the “benign” and “malignant” images in a counterfactual pair lay on opposite sides of the decision threshold (*i.e.*, they were classified as benign and malignant). In the visual-quality prescreening step, two board-certified dermatologists independently evaluated for artifacts each image that passed the classifier-consistency criterion, and we excluded images rejected by either evaluator. To ease comparison between classifiers, we included the same set of counterfactual pairs (modulo counterfactual alterations) for all classifiers; more precisely, for each reference image *x_r_*, we included the corresponding counterfactual images {*G_i_*(*x_r_*)}*_i∈C_*, where *C* represents the set of classifiers, if and only if *G_i_*(*x_r_*) passed the prescreen for each classifier *i*. For subsequent analysis, we included the 92 images from Fitzpatrick17k that passed our pre-screening criteria, and we included 100 images from ISIC to achieve a similar quantity of images.

To learn which attributes differ between benign and malignant counterfactuals—and thus influence an AI device’s predictions—we developed a two-stage annotation approach. We designed the first stage of this approach to encourage discovery of a wide variety of attributes, which we then leverage in the second stage to more efficiently collect data. Both stages leverage a graphical interface that runs locally in a web browser; expert evaluators view a pair of benign and malignant counterfactuals, then answer questions regarding (i) which member of the pair appears *most* likely to be malignant, and (ii) what attributes differ, and how they differ, between the counterfactuals. In the first stage, evaluators enter attributes as free text (*e.g.*, “skin lines more prominent”), accompanied by a “direction” specifying how the images differ (see Supplementary Fig. 8). After the first 100 pairs were evaluated by each expert, we pooled and grouped the free text terms to determine “preset” attributes (*e.g.*, “skin lines more prominent” and “more skin lines” map to the preset “Prominence of skin grooves/dermatoglyphs”) that could be selected during the second stage of annotation. This stage also retained the option for free text entry, in case a new attribute were discovered. To mitigate potential bias, we randomized and blinded evaluators to (i) the appearance order of a counterfactual pair (*i.e.* whether the benign or malignant counterfactual appeared on the left/right) and (ii) the overall order of the counterfactual pairs, including randomization of the corresponding reference images and shuffling counterfactual pairs from the various AI devices. Evaluators annotated the counterfactual pairs in sets of twenty, which required approximately 30 minutes to complete.

To infer general conclusions regarding which attributes influence the AI devices, we aggregated data from both evaluators and both stages of annotation. First, we mapped the free text attributes from the first stage of annotation to a common list of attributes, as agreed upon by the evaluators. We then filtered any counterfactual noted by either evaluator as “unable to assess” due to the presence of significant artifacts, which amounted to 4% of the total images. Finally, to obtain a global picture of each AI device, we tabulated the number of times an evaluator noted an attribute, along with the direction in which that attribute differed between the benign and malignant counterfactuals. Mathematically, we define an indicator function *s_e,c,a,d,i_* as 1 if evaluator *e* recorded for AI device *c* that attribute *a* differs in direction *d* in image *i*, and *s_e,c,a,d,i_* = 0 otherwise. Then the score for an AI device is given by the mean of *s* over images *i* ∈ *І* and evaluators *e* ∈ *ε* :

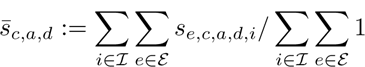

To visualize the resulting values (Fig. 2), we further aggregated the “directions” *d*, which originally included five options: *benign only*, *benign < malignant*, *different*, *benign > malignant*, and *malignant only* (during data collection, which was blinded, these terms appeared as *A only*, *A < B*, etc., where images *A* and *B* were randomized to benign or malignant). We aggregated *benign only* and *benign > malignant* into a new category, *benign*, and likewise aggregated *benign < malignant* and *malignant only* into the new category *malignant*. Finally, for each pair of AI device and classifier, we determined the “predominate direction” of that attribute, which we defined as *benign* if 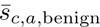 *>* 2 *·* 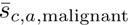, we defined as *malignant* if 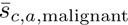 *>* 2 *·* 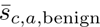, and we defined as *neither* otherwise, where the cutoff factor of 2 was chosen to prevent emphasis on small differences in frequency between the benign and malignant directions. In Fig. 2, the size of the square is then proportional to 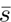 for the predominate direction, or the average of the directions if neither was predominate.

### Experimental validation of findings from counterfactuals via color shifts

To validate the attributes identified as important for dermatology AI device’s predictions in our counterfactual experiments, we aimed to experimentally modify a single attribute and observe the effect on each AI device; we chose image color as a test case, since existing mathematical tools^38^ enable well-defined, unambiguous changes to this attribute. To alter the color of each image, we converted from the sRGB color space to the CIE 1976 L*, u*, v* color space (CIELUV)^38^, added an offset to the chromaticity coordinates (*u*, v\**), then converted back to sRGB. Different chromaticity shifts were generated by varying the offset along a circle centered at (*u*, v\**) = (0, 0) with radius 20, where the factor 20 was chosen heuristically to produce color changes that we deemed visible while remaining plausible.

## Data availability

Images used in this study were obtained from publicly available repositories. ISIC images are available at https://challenge.isic-archive.com/data/. Fitzpatrick17k images are available at https://github.com/mattgroh/ fitzpatrick17k. The DDI images are available at https://stanfordaimi.azurewebsites.net/datasets/35866158-8196-48d8-87bf-50dca81df965.

Model weights for the DeepDerm classifier are available at https://zenodo.org/record/6784279#.ZFrDc9LMK-Z. The weights and model specification for the ModelDerm classifier are available at https://figshare.com/articles/ Caffemodel_files_and_Python_Examples/5406223. Model weights for our retrained variant of the SIIM-ISIC com-petition classifier are available at https://drive.google.com/drive/folders/1Zn7hNRgiI2jt7vpZO1ohpr-so9YztCCb. Scanoma and Smart Skin Cancer Detection are third party software for which we cannot redistribute model weights. At the time of writing, both are apps are available for download with no fee from the Google Play store and third-party APK package download sites.

## Code availability

Our code, including a PyTorch implementation of explanation by progressive exaggeration and classes for loading datasets and classifiers are available at https://github.com/suinleelab/derm_audit. Weights for our trained generative models and the re-trained SIIM-ISIC classifier are available at https://drive.google.com/drive/folders/1Zn7hNRgiI2jt7vpZO1ohpr-so9YztCCb.

## Author contributions

A.J.D., J.D.J., R.D., and S.-I.L. conceived of the initial study. A.J.D. prepared data and developed software for dermatology AI device reproduction, counterfactual analysis, and confirmatory experiments. A.J.D. and J.D.J. developed software for saliency map generation. Z.R.C. and R.D. analyzed counterfactual images and examined saliency maps. A.J.D., Z.R.C., J.D.J, R.D. and S.-I.L. analyzed data and designed additional experiments. Z.R.C. and R.D. provided dermatological insights and clinical context. A.J.D., Z.R.C., J.D.J., R.D., and S.-I.L. wrote the manuscript. S.-I.L. secured funding, and R.D. and S.-I.L. supervised the study.

## Data Availability

Images used in this study were obtained from publicly available repositories. ISIC images are available at https://challenge.isic-archive.com/data/. Fitzpatrick17k images are available at https://github.com/mattgroh/fitzpatrick17k. The DDI images are available at https://stanfordaimi.azurewebsites.net/datasets/35866158-8196-48d8-87bf-50dca81df965.
Model weights for the DeepDerm classifier are available at https://zenodo.org/record/6784279#.ZFrDc9LMK-Z. The weights and model specification for the ModelDerm classifier are available at https://figshare.com/articles/Caffemodel_files_and_Python_Examples/5406223. Model weights for our retrained variant of the SIIM-ISIC competition classifier are available at https://drive.google.com/drive/folders/1Zn7hNRgiI2jt7vpZO1ohpr-so9YztCCb. Scanoma and Smart Skin Cancer Detection are third party software for which we cannot redistribute model weights. At the time of writing, both are apps are available for download with no fee from the Google Play store and third-party APK package download sites.
Our code, including a PyTorch implementation of explanation by progressive exaggeration and classes for loading datasets and classifiers are available at https://github.com/suinleelab/derm_audit. Weights for our trained generative models and the re-trained SIIM-ISIC classifier are available at https://drive.google.com/drive/folders/1Zn7hNRgiI2jt7vpZO1ohpr-so9YztCCb.

## Supplementary Methods

### Saliency map generation

In initial efforts to understand the reasoning processes of the dermatology AI devices, we generated saliency maps, which highlight the regions of an image that contribute most to the AI’s prediction. To mitigate the possibility that a particular technique for saliency map generation may produce less useful results, we applied three popular techniques separately.

Following our previous work that analyzed radiology AI devices^1^, we first applied Expected Gradients^2^. This gradient-based feature attribution technique mitigates shortcomings of previous techniques^3^, including the tendency to fail to highlight darker regions of an image^4^, which would be problematic given that melanomas and melanoma look-alikes are typically darker than background skin. At a high level, this technique captures the importance of an input pixel by measuring the sensitivity of the AI devices’s prediction to small changes in that pixel (in mathematical terms, calculating the gradient), and averaging this value as the image is interpolated from a number of baseline images to the image of interest. Formally, the Expected Gradients attribution *ϕ* for a sample *x*, input feature (pixel) *i*, baseline distribution *D*, and AI device *f* is given by:

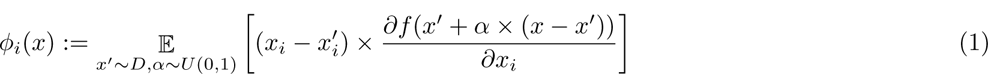

As our background distribution, we chose the full ISIC 2019 dataset; attributions were estimated via Monte Carlo sampling, using 1000 samples.

As our second feature attribution technique, we next calculated saliency maps via KernelSHAP^5^. This technique characterizes the importance of an input pixel by measuring how the model’s prediction changes when that feature is “removed” (in our case, replaced by the mean color of that image). Importantly, feature interactions are properly accounted by removing multiple features at a time, then summarizing a feature’s effects over these subsets via the *Shapley value*^6^, a well-established technique grounded theoretically in game theory. KernelSHAP estimates the Shapley value by casting it as the solution to a least squares problem, which can be solved by sampling random sets of features to remove, rather than requiring exhaustive enumeration of every possible set of features. To enable tractable calculation, we define 16 *×* 16 super-pixels as features, then upsample the final result via bilinear interpolation to match the original image size. For each image, we perform the KernelSHAP estimate using 10^5^ samples, which required approximately one hour of computation on an NVIDIA RTX 2080Ti graphics processing unit, per image.

Finally, we calculated saliency maps via the highly popular GradCAM approach^7^. This technique characterizes the importance of a region of an image by monitoring the activation of individual neurons in a neural network, which may retain coarse spatial information even in layers far from the input. Specifically, for each channel of an activation map, the technique multiplies that activation by the derivative of the network’s output with respect to that neuron, then sums over all channels to determine an aggregate value for a spatial location, before finally discarding negative values. Formally, let *A* denote the activations of a neural network *f* at the layer of interest, let *k* represent each channel of those activations, and let *x* denote the input. Then the GradCAM attributions *ϕ* are given by:

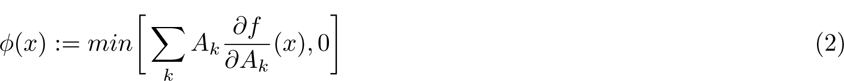

We take derivatives of the model’s prediction of the likelihood of melanoma such that intuitively, these attributions can be understood as identifying the regions of the image that contribute toward a prediction of melanoma. As the “layer of interest,” we target the layer immediately prior to the final global pooling. To account for model ensembling in the AI device SIIM-ISIC, which includes three individual models, each of which is evaluated at test time on eight versions of the input image (the original, plus a series of flips and rotations), we treat the channels of that layer of the twenty-four resulting “sub-models” as channels of one aggregated layer. In other words, in the above equation, *k* runs over the channels of each sub-model’s final layer prior to global pooling, as well as over all sub-models; to preserve spatial relationships, we reverse the augmentations before averaging. The resulting saliency maps match the spatial dimensions of the layer of interest, which (as is typical with GradCAM) are lower resolution than the input image; we upsample the saliency map via a bilinear filter to match the dimensions of the original image.

In all cases, we display the final saliency map by taking its absolute value, then overlaying it on a desaturated version of the original image, with the saliency map blended at α = 80%. To mitigate overemphasis of the color scale on outlier values, we clip the maximum value of the saliency map at the 99*^th^* percentile of each image.

### Supplementary Tables

**Supplementary Table 1.** | Reference for attribute names from main text Fig. 2, which for conciseness shortens the original attribute names used during our annotation procedure. Some attributes were not shortened: “atypical pigment networks”, “blue/white veil”, and “erythema” (ISIC); “erythema”, “telangiectasia”, and “uneven pigmentation” (Fitzpatrick17k).

|  | Category | Shortened | Original |
| --- | --- | --- | --- |
| ISIC | lesion | darker pigmentation | color of pigmentation - darker |
|  | lesion | number of colors | number of colors in lesion |
|  | background | pinker | color - pink |
|  | lesion | structureless areas | structureless area(s) |
|  | background | brown spots | number or prominence of brown spots |
|  | background | hair | number or prominence of hairs |
|  | lesion | regression | prominence of regression |
| Fitzpatrick17k | lesion | darker pigmentation | color of pigmentation - darker |
|  | background | darker pigmentation | color of pigmentation - darker |
|  | lesion | number of colors | number of colors in lesion |
|  | lesion | erythema | redness/erythema |
|  | background | hair | prominence of hair |
|  | lesion | browner pigmentation | color of pigmentation - brown |
|  | lesion | scale/crust | presence of scale/crust |

**Supplementary Table 2.** | Kernel inception distances (×10^−3^) between generated images and the reference dataset. The reference dataset contains all images from ISIC or Fitzpatrick17k, after exclusions (see Methods: Image selection and preprocessing), i.e., it was not limited to those images evaluated by experts. The generated dataset contains, for each image in the reference dataset, either the “benign” or “malignant” counterfactual chosen uniformly at random.

|  | DeepDerm | ModelDerm | Scanoma | SSCD | SIIM-ISIC |
| --- | --- | --- | --- | --- | --- |
| ISIC | 13.1 | 19.5 | 9.6 | 16.1 | 16.0 |
| Fitzpatrick17k | 22.7 | 19.6 | 23.0 | 8.8 | 23.6 |

### Supplementary Figures

**Supplementary Fig. 1.**
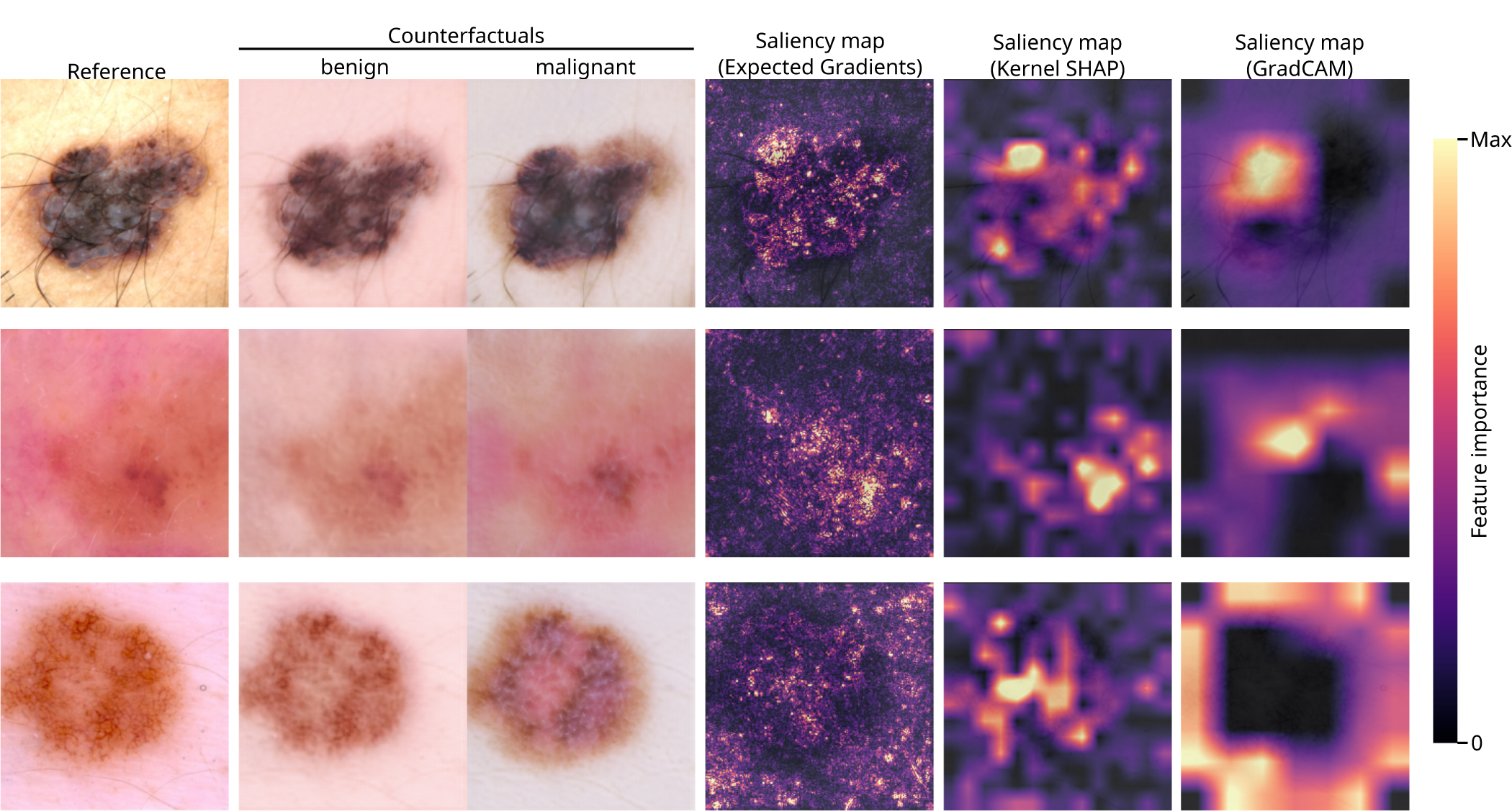
| Comparison of insights from counterfactuals and saliency maps. We calculated feature attributions using three popular techniques, Expected Gradients^2^, Kernel SHAP^5^, and GradCAM^7^ (see Supplementary Methods) and then produced our best-effort visualizations of the resulting saliency maps. We failed to gather insights from the saliency maps, except that the AI device may focus on the lesion (but perhaps not always, depending on the saliency technique). In contrast, the counterfactuals provided more granular and medically interpretable insights: for instance, based on the malignant counterfactuals we inferred that multiple colors of pigment (top + bottom), erythema (middle + bottom), darker pigmentation (all), and blue-white veil (bottom) tend to elicit more malignant predictions. In this figure, all saliency maps and counterfactuals were generated in reference to our AI device “SIIM-ISIC”.

**Supplementary Fig. 2.**
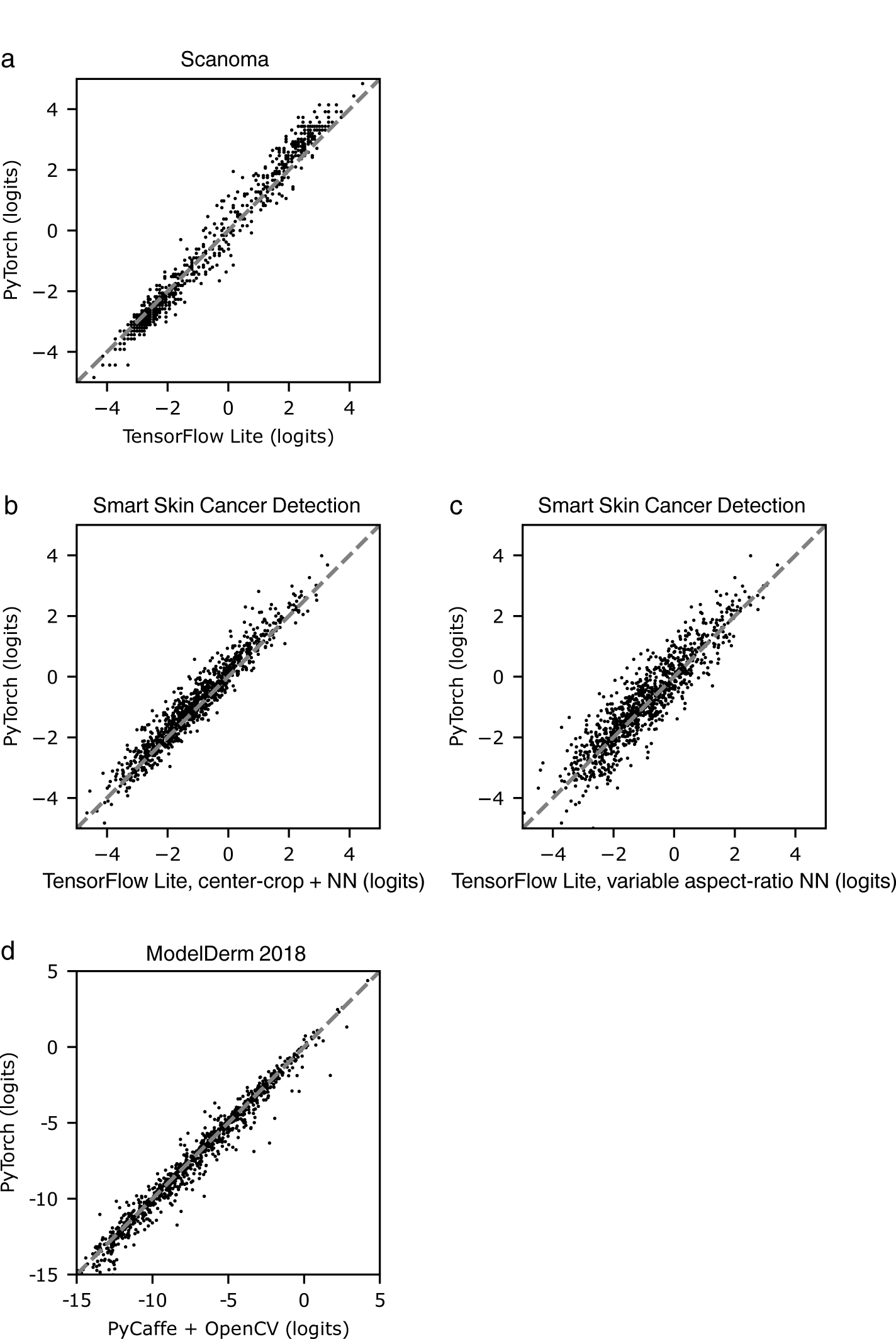
| Similarity of predictions from original classifiers and our PyTorch re-implementations. To evaluate our PyTorch re-implementations’ similarities to the original models, we compared the classifiers’ predictions on a series of 1000 images from the ISIC dataset. **a**, Comparison of our PyTorch re-implementation of Scanoma with a TensorFlow Lite implementation of Scanoma, which differs from the original Android implementation only by antialiasing constants in the bilinear filtering preprocessing step. We compared our PyTorch re-implemention of Smart Skin Cancer Detection (SSCD), which uses square center-cropping and bilinear resizing to preprocess images, against the original TensorFlow Lite implementation with square center-cropping and nearest-neighbor resizing (**b**), and nearest-neighbor resampling to a square input (allowing changes to the aspect ratio, **c**). Aside from the input resizing routine, our PyTorch implementation achieves identical outputs to the original TensorFlow Lite classifier. **d**, Comparison of our PyTorch reimplemention of ModelDerm 2018, including our differentiable histogram equalization preprocessing step, with the original PyCaffe and OpenCV implementation. NN, nearest-neighbor interpolation.

**Supplementary Fig. 3.**
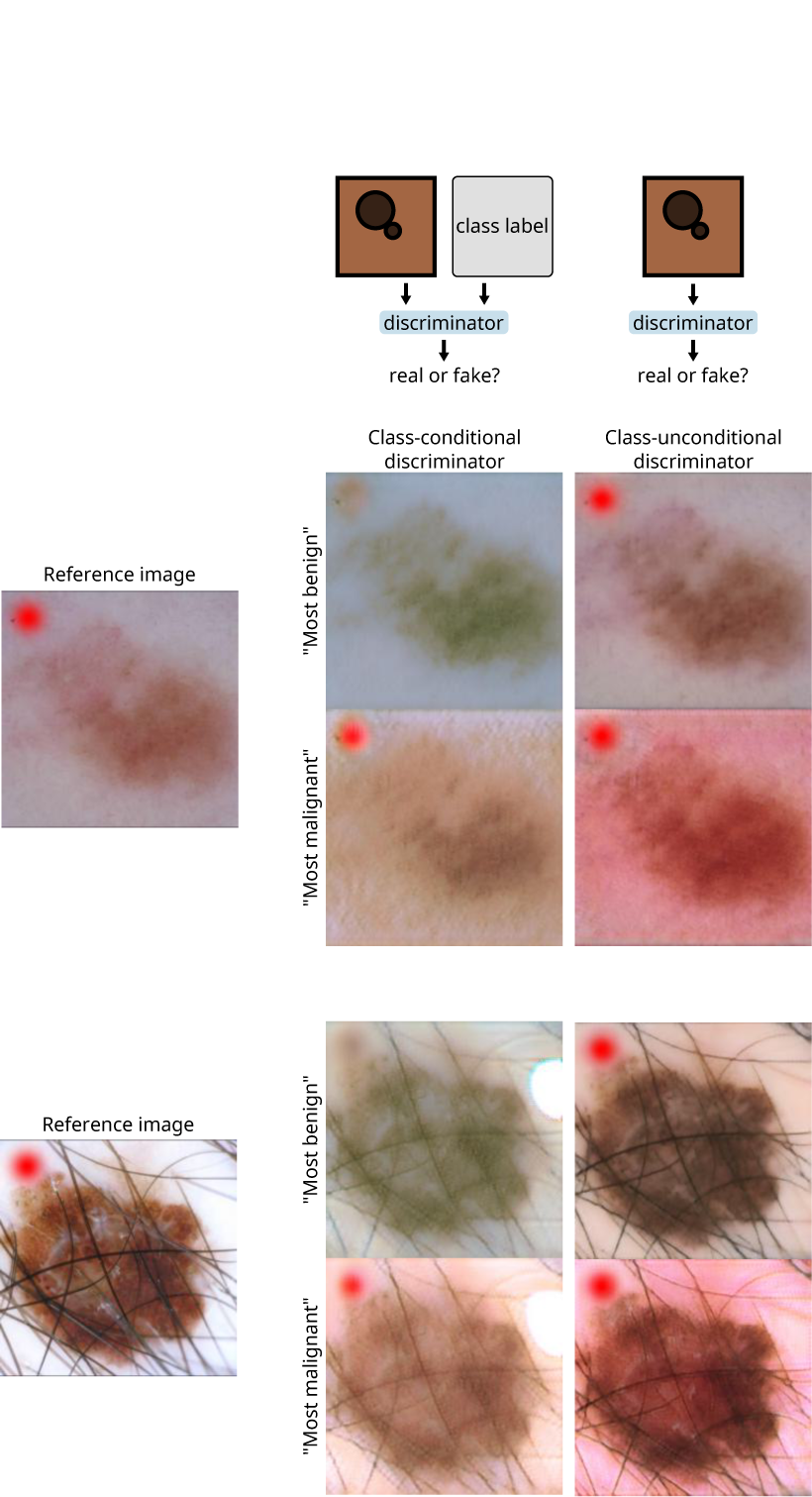
| Comparison of a class-conditional discriminator with a discriminator not conditioned on class, with respect to their treatment of features correlated with the classifier’s output. Hypothesizing that a class-conditional discriminator would alter features correlated with a classifier’s predictions, even when not used by the classifier, we designed a scenario in which the classifier is unlikely to depend on the presence of a test artifact (a red dot in the corner of the image), but the test artifact correlates with melanoma status in the training data for the generative model. In particular, we trained an EfficientNet-B7 to detect benign versus malignant lesions among the melanomas and melanoma-lookalikes of the ISIC 2019 training data; since this training data lacked the test artifact in any image, the classifier is unlikely to depend strongly on the presence of the artifact. When training the generative models, we introduced the test artifact into every image of a melanoma, such that it correlates perfectly with melanoma status. While the test artifact is altered by the generator that was trained in conjunction with the class-conditional discriminator, which could mislead an investigator to conclude that the classifier’s prediction is based in part on the presence of the test artifact, the generator trained with the discriminator that is not conditioned on class leaves the test artifact unaltered. In addition, we anecdotally noted that the generator trained with the unconditional generator produced images of higher visual quality (bottom two rows).

**Supplementary Fig. 4.**
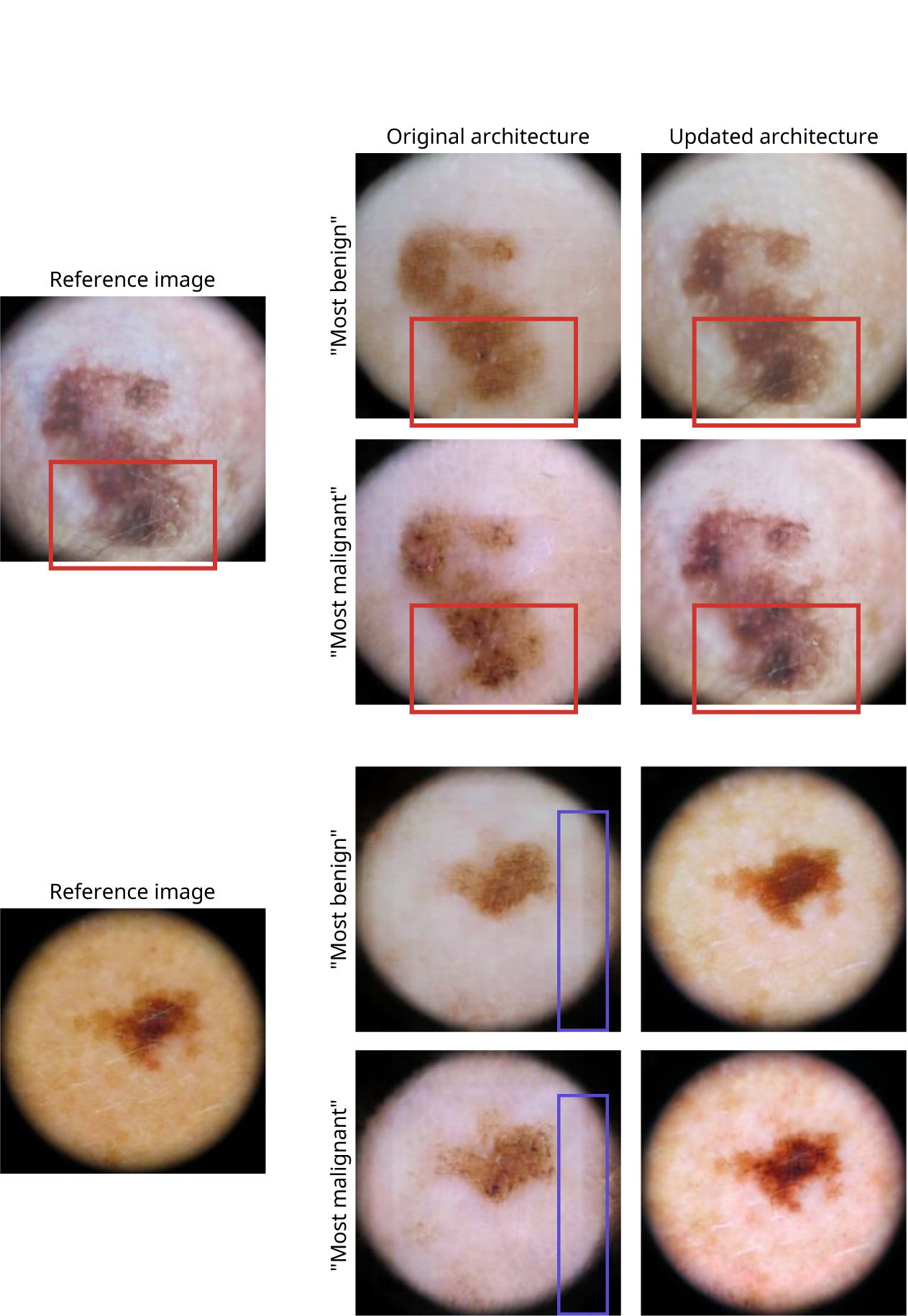
| Comparison of the visual quality of images produced by the original generator architecture from ref.^8^ with those produced by our updated architecture. Our updated architecture successfully reproduces details such as hairs, which the original architecture fails to capture (red boxes). The original architecture also introduces linear artifacts (blue boxes) not present in the original image, while we noted no such artifacts in images generated by the updated architecture.

**Supplementary Fig. 5.**
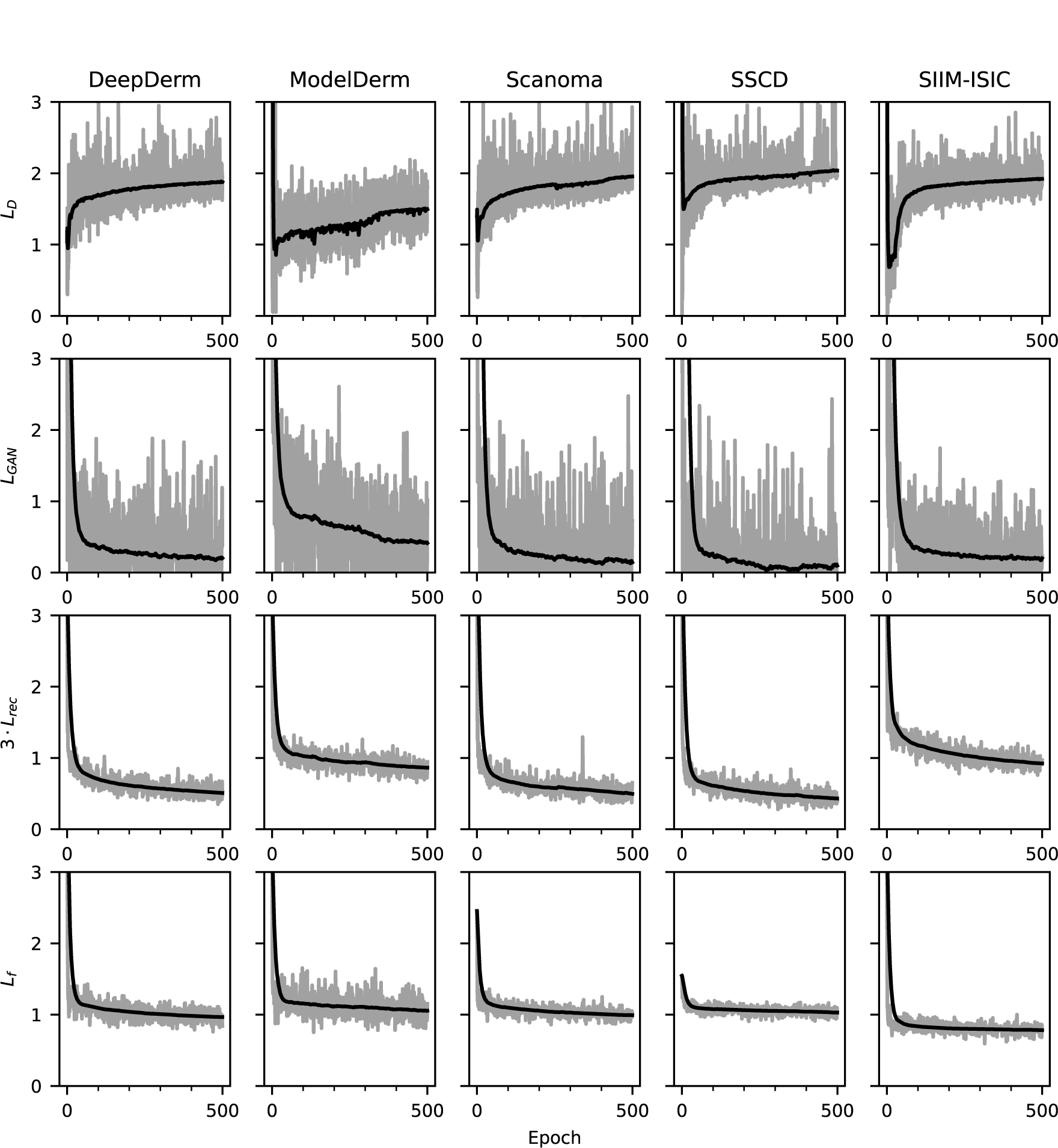
| Evolution of loss terms during training of our generative models on the ISIC dataset. Loss terms are plotted after multiplication by their respective scaling factors (*λ_rec_* = 3, *λ_D_* = *λ_GAN_* = *λ_f_* = 1). Gray lines indicate the instantaneous loss, and black lines indicate the exponential moving average (*α* = 0.001; loss terms were recorded at each gradient update of their respective model).

**Supplementary Fig. 6.**
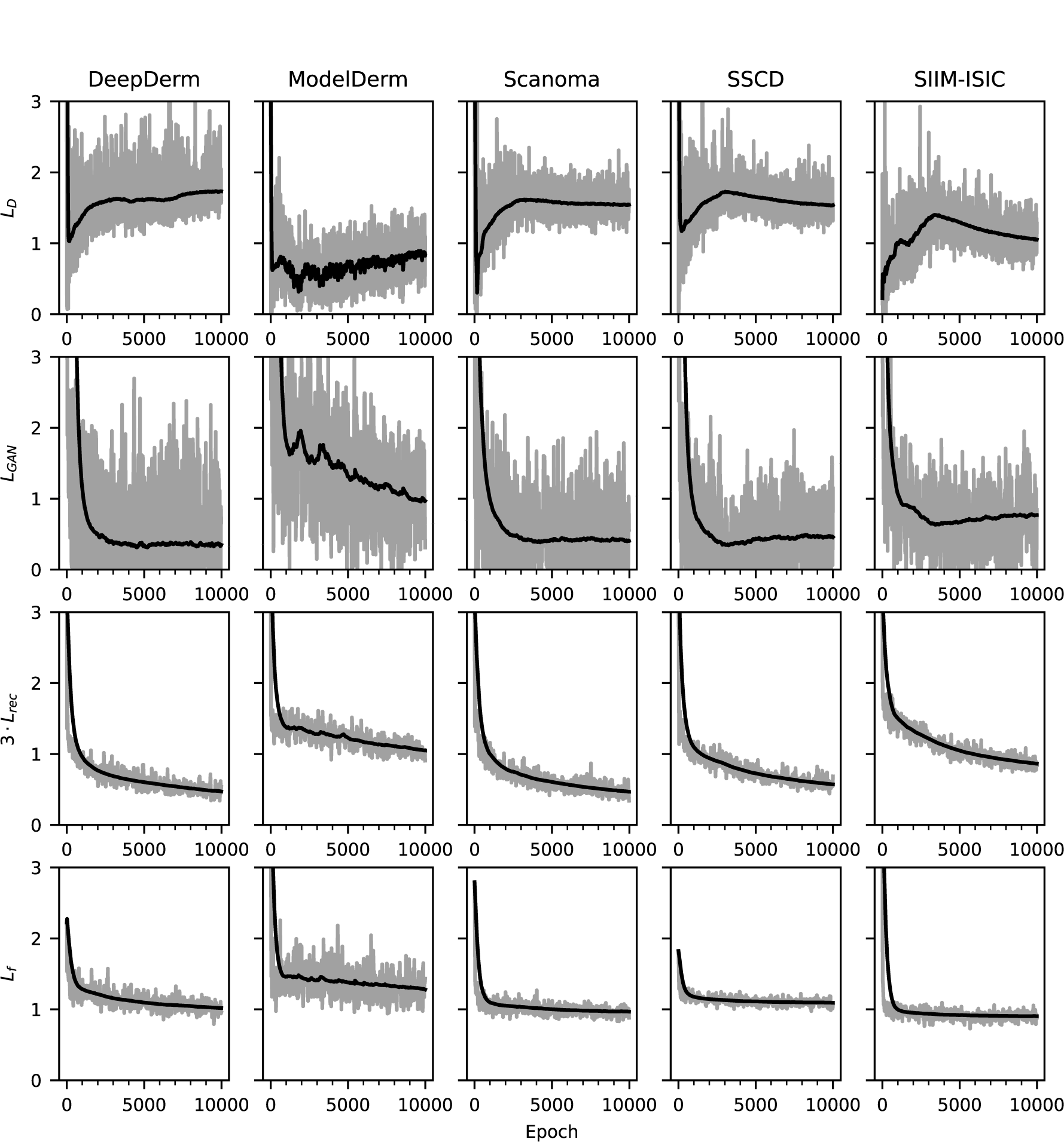
| Evolution of loss terms during training of our generative models on the Fitzpatrick17k dataset. Loss terms are plotted after multiplication by their respective scaling factors (*λ_rec_* = 3, *λ_D_* = *λ_GAN_* = *λ_f_* = 1). Gray lines indicate the instantaneous loss, and black lines indicate the exponential moving average (*α* = 0.001; loss terms were recorded at each gradient update of their respective model).

**Supplementary Fig. 7.**
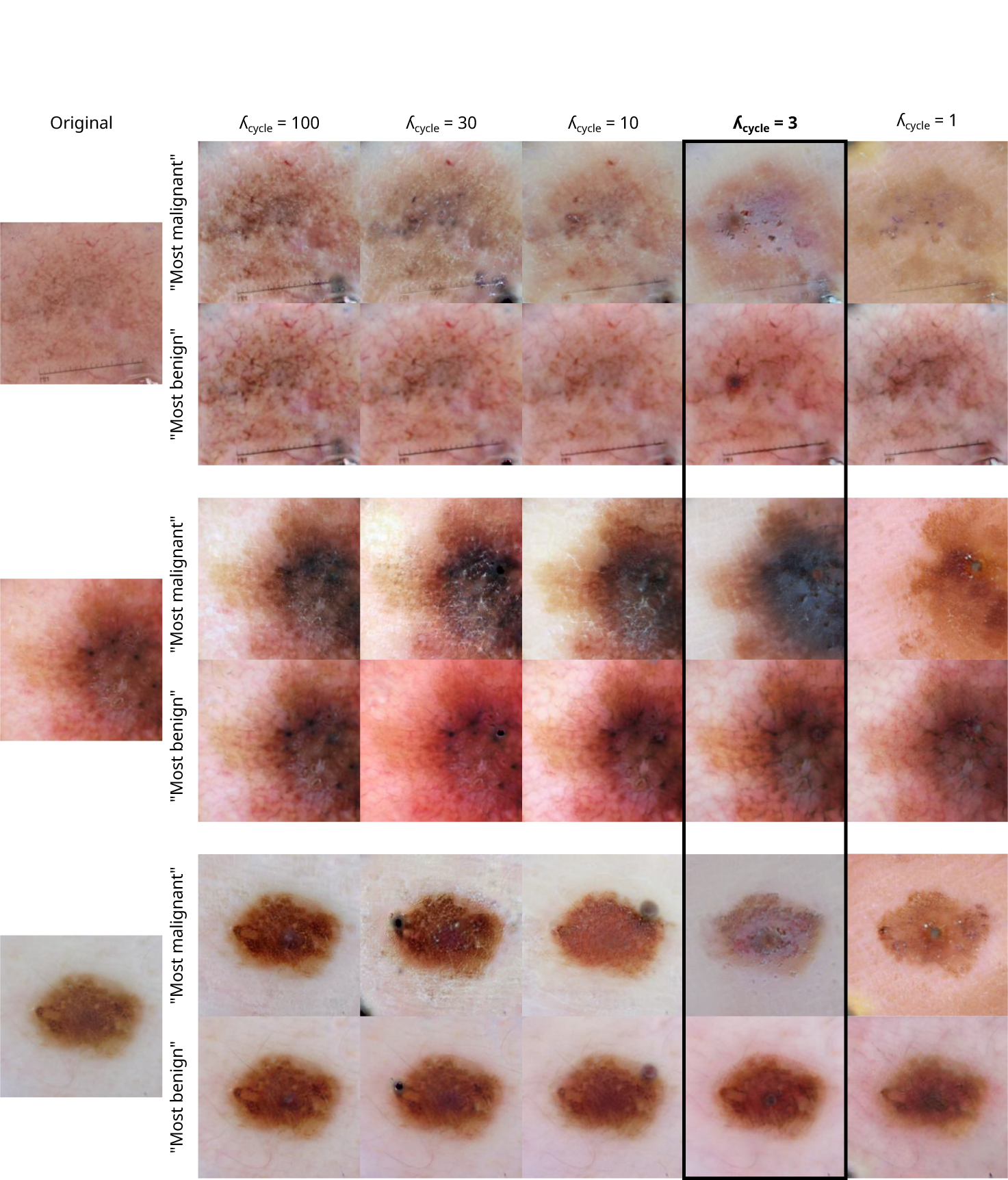
| Tuning of the hyperparameter λ_cyc_ in the generative models. To tune the hyperparameter *λ_cyc_*, we started with the value of 100 reported in the original publication of Explanation by Progressive Exaggeration^8^, then progressively decreased its value until the alterations between the “most benign” and “most malignant” images became apparent (based on manual, visual inspection), while ensuring that the generated images still appeared similar to the original, reference image. Counterfactuals in this figure were generated to analyze the AI device ModelDerm; images were chosen uniformly at random.

**Supplementary Fig. 8.**
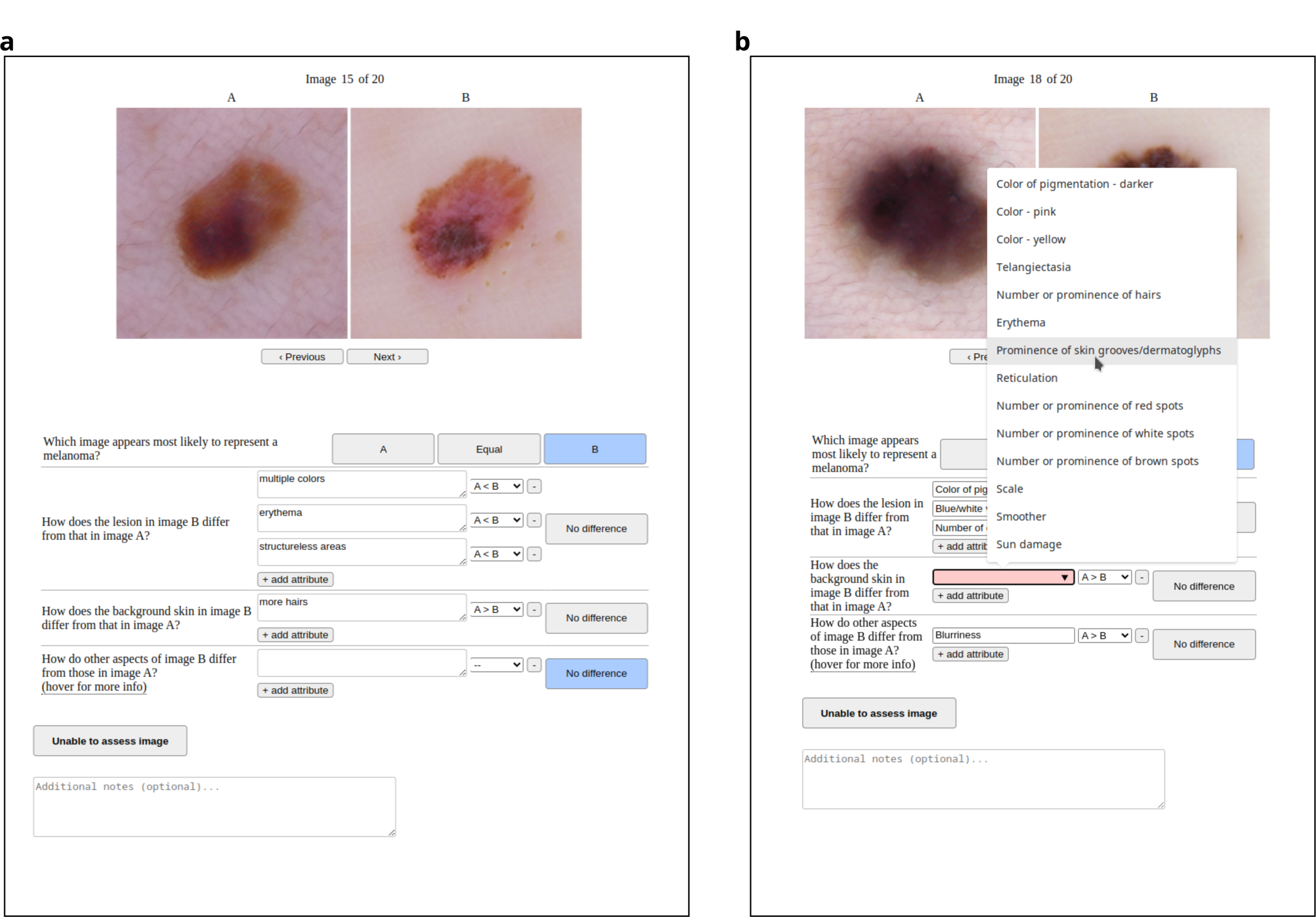
| Screenshots of app for expert analysis of counterfactuals. **a**, “Free text” version of the app, used during the initial phase of data collection to encourage collection of a broad, diverse set of attributes that differ between benign and malignant counterfactuals. The expert annotator enters an attribute (*e.g.*, “structureless areas”) and then specifies how that attribute differs between the two images by selecting a comparator (“A only”, “A *>* B”, “A *<* B”, “B only”, or “different”) from the drop down menu. The app allows entry of an arbitrary number of attributes, and contains multiple categories of attributes (“lesion”, “background”, and “other”) to remind annotators to pay attention to each part of the counterfactuals. **b**, After the initial phase of free-text data collection, attributes are pooled and grouped in collaboration with the expert annotators, to produce a list of “preset” responses that enables faster, more uniform analysis. In the remaining modules, expert annotators may select a preset from a drop-down list, or continue to enter attributes as free text, accounting for the possibility that new attributes are discovered after the initial free-text phase.

**Supplementary Fig. 9.**
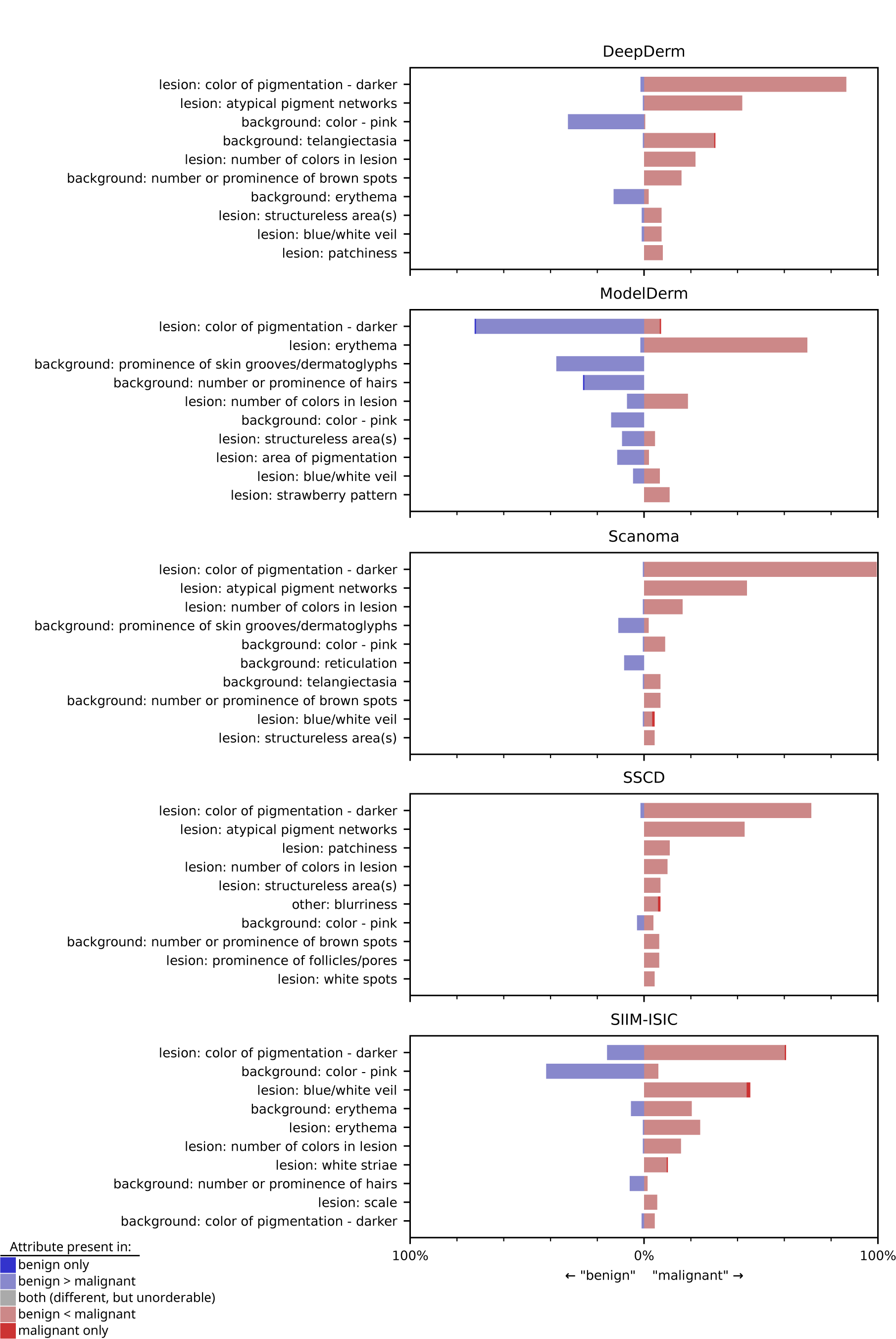
| Attributes identified by our join expert-XAI auditing procedure as key influences on the output of individual dermatology AI devices, when evaluated on the ISIC dataset. In contrast to main text Fig. 2, attributes are ordered by the proportion of counterfactual pairs *from the specified AI device* in which experts noted that attribute differs, enabling examination of attributes relevant to a particular AI device but not necessarily to most AI devices (*e.g.*, prominence of skin grooves or dermatoglyphs, which influences Scanoma and ModelDerm).

**Supplementary Fig. 10.**
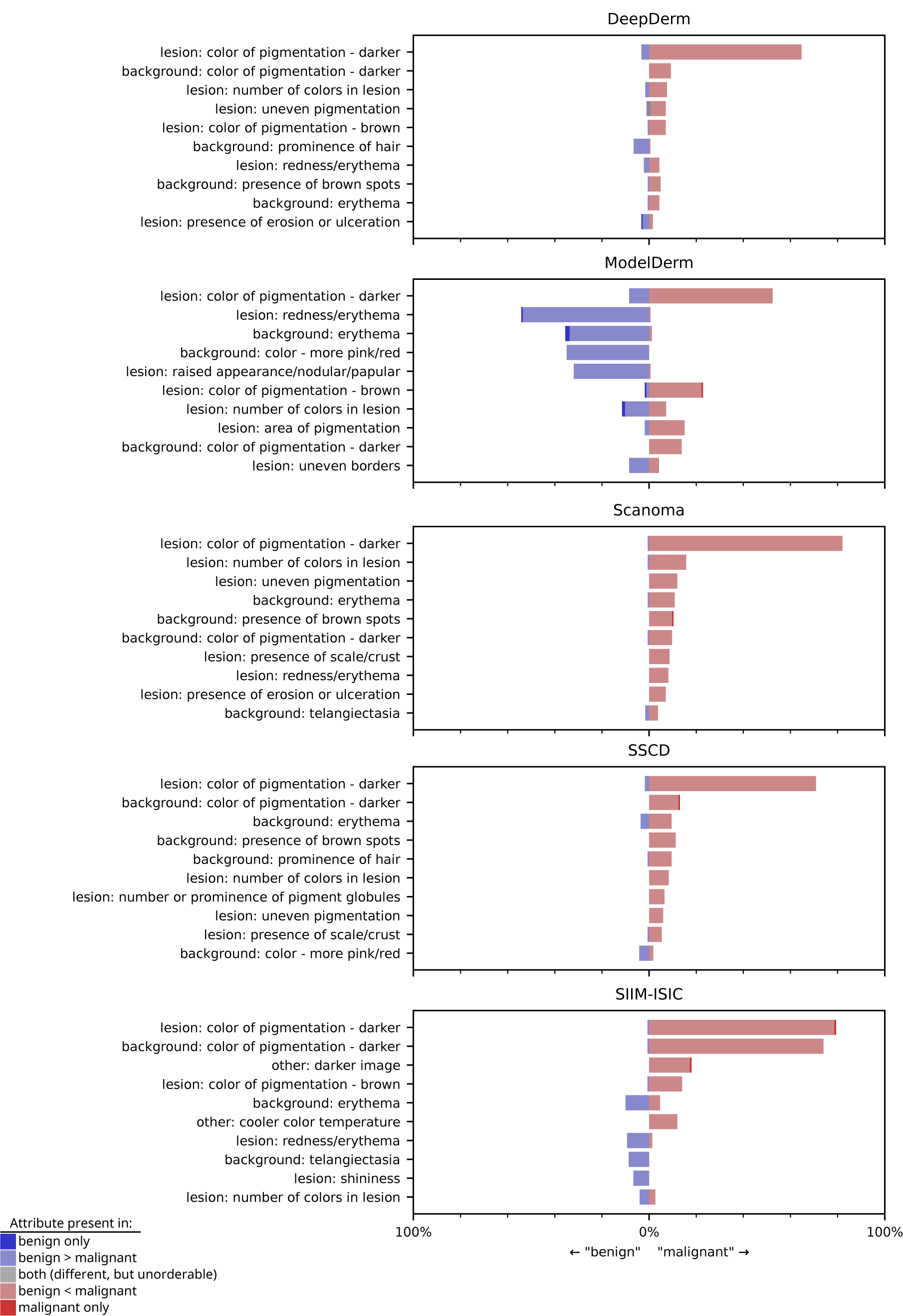
| Attributes identified by our join expert-XAI auditing procedure as key influences on the output of individual dermatology AI devices, when evaluated on the Fitzpatrick17k dataset. In contrast to main text Fig. 2, attributes are ordered by the proportion of counterfactual pairs *from the specified AI device* in which experts noted that attribute differs, enabling examination of attributes relevant to a particular AI device but not necessarily to other AI devices.

**Supplementary Fig. 11.**
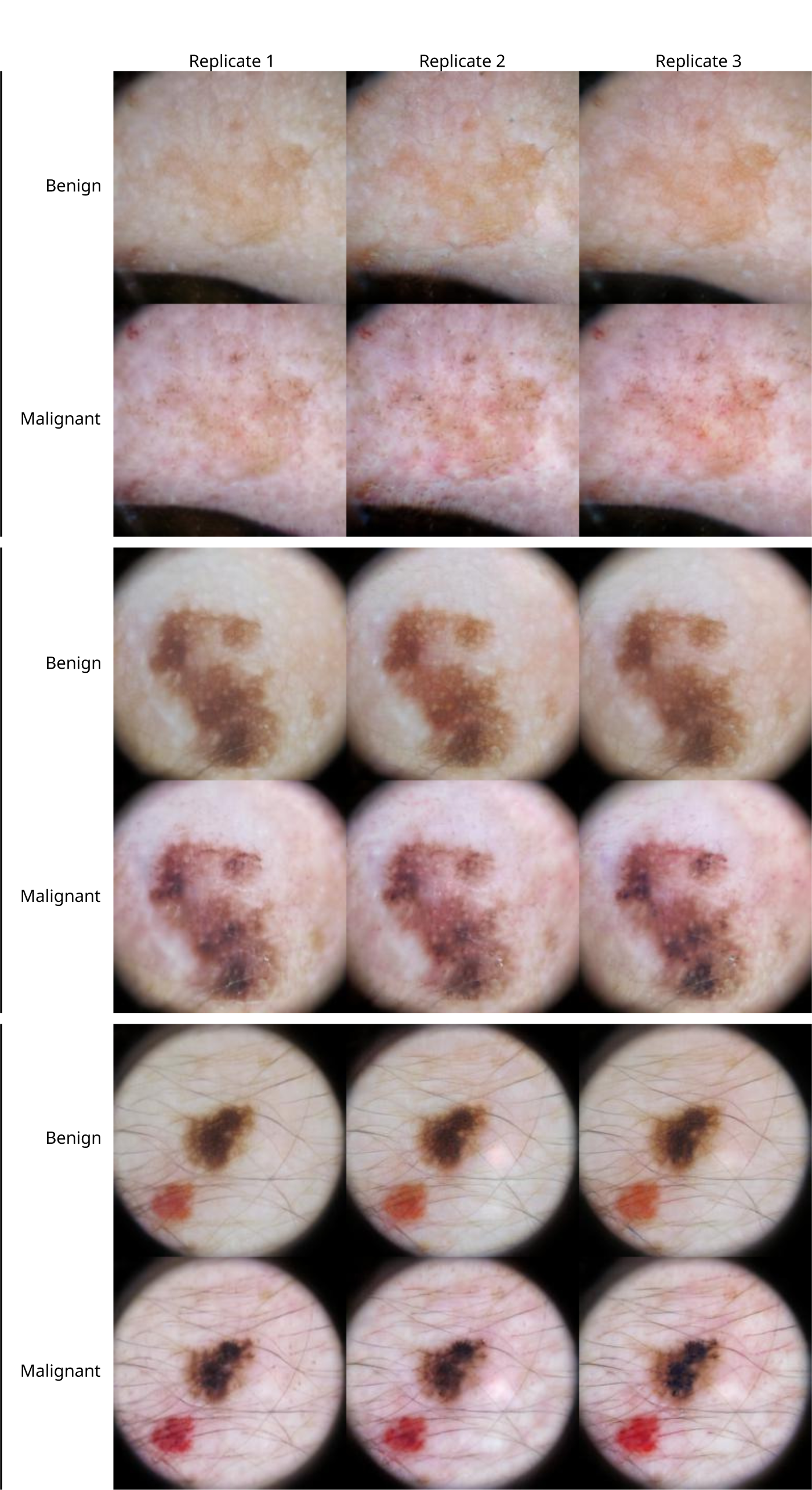
| Independent re-trainings of a generative model using the same training data and AI device. Retrainings preserve key attributes that vary between benign and malignant counterfactuals, such as erythema of the background skin (top), darker pigmentation of the lesion (middle), and multiple colors of pigment in the lesion (bottom). The generative models were trained to evaluate the AI device Scanoma. Images are adapted with permission from the ISIC dataset^9–11^.

